# Digital Treatment Signatures: Cardiovascular Risk Prediction Using Antihypertensive Medication Fill History

**DOI:** 10.64898/2026.09.03.26362175

**Authors:** Yifan Zheng, Karen B. Farris, Antoinette B. Coe, Michael P. Dorsch, Kayvan Najarian, Corey A. Lester

**Affiliations:** Department of Clinical Pharmacy Translational Science, University of Michigan College of Pharmacy, Ann Arbor, MI, USA; Department of Computational Medicine and Bioinformatics, University of Michigan, Ann Arbor, MI, USA; Max Harry Weil Institute for Critical Care Research and Innovation, University of Michigan, Ann Arbor, MI, USA; Michigan Institute for Data and AI in Society (MIDAS), University of Michigan, Ann Arbor, MI, USA; Center for Data-Driven Drug Development and Treatment Assessment, University of Michigan, Ann Arbor, MI, USA

**Keywords:** medication adherence, antihypertensive medications, cardiovascular diseases, risk assessment, electronic health records

## Abstract

**Objective:** Taking a blood pressure medication is not a single fact but a trajectory of fills, gaps, and regimen changes. Dispensing records capture that trajectory, yet risk calculators reduce it to a yes/no treatment indicator and quality programs to proportion of days covered (PDC). We asked how much of that discarded signal is recoverable.

**Materials and Methods:** We studied 7,625 adults treated for hypertension using linked electronic health record and pharmacy dispensing data. From a 1-year medication-history window, we predicted 3-year myocardial infarction, stroke, or all-cause death. Holding cohort, covariates, horizon, and validation fixed, we compared PDC, engineered temporal and regimen features, and the ordered fill sequence modeled with an attention-based bidirectional LSTM survival network. Factorial analyses separated representation from model class.

**Results:** C-index increased from 0.7113 with clinical factors alone to 0.7131 with PDC, 0.7273 with engineered summaries, and 0.7396 with the ordered sequence (ΔC=0.0283; 95% CI, 0.0189–0.0382). The sequence exceeded engineered summaries by 0.0123 (0.0040–0.0202). Richer representations improved discrimination by 0.0181–0.0191, approximately three times the 0.0059–0.0078 from changing model class. Among patients with PDC ≥0.80, PDC ranked risk poorly (C=0.4628), whereas sequence-model strata had 3-year event rates of 3.6%–22.8%.

**Discussion:** A medication history’s prognostic value lies in its temporal structure, which aggregate adherence measures discard, so the sequence advantage reflects preserved information rather than model complexity. External validation is needed.

**Conclusion:** Traditional metrics of antihypertensive treatment conceal substantial risk heterogeneity, whereas a tokenized dispensing timeline recovers a prognostic treatment signature that improves cardiovascular risk prediction from existing records.

## Background and Significance

Antihypertensive medication use is inherently longitudinal, yet cardiovascular risk prediction and pharmacy quality measurement typically reduce that history to highly compressed summaries. Electronic health records primarily document what clinicians prescribe rather than what patients obtain, whereas pharmacy dispensing data capture the timing and continuity of medication acquisition. Despite that richer information, standard approaches usually represent antihypertensive use in one of two ways. Cardiovascular risk calculators, including the Pooled Cohort Equations[1] and PREVENT equations[2], use a binary indicator of whether a patient is receiving antihypertensive treatment. Pharmacy quality programs, including the Medicare Star Ratings, summarize dispensing history using the proportion of days covered (PDC)[3]. PDC informs health-plan rankings and payment, has been associated with cardiovascular outcomes[4,5], and helps identify patients for adherence interventions. Both representations discard potentially informative features of medication use, including refill timing, interruptions, regularity, and regimen changes.

This compression may be appropriate for the purposes for which these measures were developed, but it is not clear that it preserves information relevant to individual prognosis. The treatment indicator is designed primarily to account for the fact that observed blood pressure may already reflect treatment; among patients who are all receiving therapy, however, it is constant and therefore provides no discrimination. PDC serves a different purpose: it provides a standardized measure of medication coverage for quality assessment and adherence monitoring. In the Medicare Star Ratings, a threshold of 0.80 is commonly used to classify patients as adherent[3]. Once medication use is summarized in this way, substantial heterogeneity in the timing and structure of medication use may remain unrepresented. That heterogeneity is also socially patterned: antihypertensive nonadherence measured by PDC is more common among Black and Hispanic patients and in lower-income groups[6].

Prior studies have established that medication adherence is associated with cardiovascular outcomes and that longitudinal adherence trajectories may contain additional outcome information[7–9]. However, association does not necessarily imply improved prognostic discrimination beyond established clinical predictors such as age, prior cardiovascular disease, kidney disease, and metabolic risk factors[10,11]. Moreover, observed associations between adherence and outcomes may partly reflect behavioral or clinical characteristics captured by the healthy-adherer effect rather than medication coverage alone[12–14]. The more important unresolved question, therefore, is how much prognostic information is lost when a longitudinal medication history is compressed and whether that information can be recovered.

Existing studies have generally evaluated adherence as an additional covariate or compared alternative longitudinal models without isolating the contribution of data representation from that of model flexibility. As a result, it remains unclear whether improved prediction arises because richer medication histories contain additional prognostic information or simply because more complex algorithms fit the data more effectively. We therefore evaluated antihypertensive medication history as a hierarchy of representations, from conventional summary measures to engineered temporal features and the ordered dispensing sequence itself, while holding the cohort, clinical covariates, outcome, prediction horizon, and validation framework constant. We further examined whether richer representations distinguish cardiovascular risk among patients already classified as adherent by conventional PDC thresholds.

## Objective

We evaluated how the representation of longitudinal antihypertensive dispensing history affects cardiovascular risk prediction. Holding the cohort, outcome, clinical covariates, prediction horizon, and validation framework constant, we compared average PDC, engineered longitudinal features, and the ordered sequence of individual fills. We further explored how richer representations improved risk discrimination beyond conventional PDC and among patients already classified as adherent.

## Materials and Methods

### Study design and setting

We conducted a retrospective cohort study of adults treated with antihypertensive medications at Michigan Medicine, an academic health system, linking electronic health records to outpatient pharmacy dispensing records obtained through Surescripts, a national electronic-prescribing and medication-history network[15]. The study followed the Declaration of Helsinki and was approved by the University of Michigan Medical School Institutional Review Board (HUM00244597), which waived informed consent because the research uses existing data collected in the course of clinical care. Reporting follows TRIPOD+AI[16], and the analysis plan was registered internally before model fitting.

### Study population

Patients entered the cohort at their first observed antihypertensive fill that was preceded by a clinical encounter within the prior year. Because dispensing is observed only from the start of the linked pharmacy data, the cohort comprises prevalent rather than new users. We additionally required at least two antihypertensive fills and age 18 years or older. Patients entered between June 2010 and August 2019, and deaths were ascertained from the state death index through November 2025. Each medication was assigned to one of seven antihypertensive classes.

Baseline clinical covariates were taken from the electronic health record as the most recent value on or before the index date: age, sex, race, systolic blood pressure, total and HDL cholesterol, kidney function estimated with the race-free 2021 CKD-EPI creatinine equation[17,18], smoking status, and body-mass index. Race is used as a sociopolitical category recorded in the record, not as a measure of genetic ancestry. With prior cardiovascular disease, the remaining comorbidity block, and prior-year encounter count, these form the 22-covariate specification shared by every model.

### Outcome

The outcome was a major adverse cardiovascular event (MACE): the first myocardial infarction, stroke, or death from any cause after the index date. This follows the conventional three-point structure[19] but substitutes all-cause for cardiovascular death, which is completely ascertained during follow-up and leaves the composite free of competing risks. Heart failure was recorded separately and not included.

### Representations of the medication history

All models included the baseline clinical covariates. To these we added progressively more detailed representations of antihypertensive history, ordered to reflect and then extend current practice.

The reference model used the clinical covariates alone, which is how risk calculators such as the Pooled Cohort Equations and PREVENT[1,2] represent antihypertensive therapy, as a single on-treatment indicator. The next model added the proportion of days covered, the adherence measure used in the Medicare Star Ratings[3]. Subsequent models added the drug classes a patient was taking and then measures of regimen change, including new class additions, class switches, and refill frequency; the full engineered summary combined all of these.

Each summary was cumulative, reflecting the complete fill history up to any given time, so that the summary models and the sequence model drew on the same information. Adherence was the proportion of days on which any antihypertensive supply was available, with overlapping coverage counted once. Time-varying features accumulated on a 30-day interval scaffold and used only information available before the prediction time. They comprised cumulative new-class additions, class switches, refill rate, distinct-class count, dose intensity, and the longest coverage gap.

A final model used no engineered summaries. It read each fill in sequence, described only by information available at dispensing: drug class, time since the index date, time since the previous fill, days supplied, dose intensity, and whether the fill was early. A bidirectional long short-term memory (LSTM) network with an attention layer summarized the sequence[20–22]. Features that duplicated the summary models were excluded, so that any difference reflected the ordering and timing of fills.

Each fill was encoded as a fourteen-dimensional token, and the network was trained on the Cox partial likelihood with Efron’s approximation for tied event times[23]. Fitting used 20 imputations by 5 folds by 3 random seeds, and each patient’s held-out risk score was averaged over the three seeds. Architecture and optimization detail is given in Supplementary Methods.

### Missing data

Five measurements were incompletely recorded: systolic blood pressure (41%), body-mass index (30%), and total cholesterol, HDL cholesterol, and creatinine (13% to 17%). We generated 20 imputed datasets[24,25] and recalculated kidney function from imputed creatinine[17,18]. Imputation models were fitted once on the full cohort rather than within each cross-validation fold and included the outcome, which may have inflated absolute discrimination; every representation is built on the identical imputed covariates, and the medication inputs are never imputed. Results were combined using Rubin’s rules[26].

### Statistical analysis

All summary models estimated time-to-event risk using Cox proportional-hazards regression[27], and the neural-network model was trained with the same survival objective. Prediction was made at a fixed landmark one year after the index date[28], using the history accumulated up to that landmark to estimate the risk of MACE over the following three years. The Cox models were estimated on summary values frozen at the landmark against post-landmark follow-up, the same task on which the sequence model was trained.

The primary, prespecified measure was discrimination, quantified with a time-dependent concordance statistic where 0.5 indicates chance[29,30]. Performance was estimated by five-fold cross-validation across the 20 imputed datasets, with the neural-network model averaged over three training runs, so that all reported metrics derive from held-out predictions. The analysis plan gave equal weight to calibration and to clinical usefulness, assessed by net benefit in decision-curve analysis[31] at thresholds of 7.5%, 15%, and 25%[1].

Antolini’s time-dependent concordance[32] was the prespecified secondary measure; anchored at the landmark, it uses all post-landmark events rather than only those in a single three-year window. Differences between models were tested with a paired bootstrap comparing models on the same resampled patients, computed on the set of patients with valid predictions from every model so that all share one denominator. The proportional-hazards assumption was not formally assessed, and no hazard ratio from the ladder is interpreted.

Because models that rank patients well may still misstate absolute risk, we prespecified recalibration of the neural-network model by leave-one-fold-out fitting within each imputed dataset, comparing a scaling correction[33] with a more flexible monotonic correction[34]. Recalibration does not reorder patients, so discrimination is unchanged. Absolute risks were obtained by calibrating each score to the observed survival function.

Analyses were performed in Python, with lifelines, XGBoost under its Cox objective, PyTorch, scikit-learn, and miceforest. Package versions are recorded in the environment specification included with the analysis code.

### Secondary and sensitivity analyses

To separate the contribution of the representation from that of the model, we refit each summary representation with a nonlinear gradient-boosted survival model[35] and compared the gain from enriching the representation with the gain from changing model class (Supplementary Table S3). We also fit an unordered variant of the sequence model, a model using only counts of fills and encounters, and one adding the raw fill count to the full engineered summary (Supplementary Table S4).

To examine whether average adherence obscures within-patient heterogeneity, we stratified patients into three bands: PDC of 0.80 or above, the band the Star Ratings certify as adherent, 0.50 up to 0.80, and below 0.50. Within each band we computed the concordance of average adherence and of the sequence model, and the observed three-year incidence of MACE across thirds of sequence-model predicted risk. Because the sequence model also carries the clinical covariates, we scored three design-matched references on the same patients: the covariates alone, the covariates plus average adherence, and the fill sequence with no covariates.

Because the primary composite included death from any cause, we assessed whether the discrimination attributable to the dispensing sequence was specific to mortality. Using the underlying cause of death from the state death index, we defined a cardiovascular outcome as the first myocardial infarction, stroke, or death with an underlying cause in ICD-10 chapter I, censored non-cardiovascular deaths, and refitted every representation against it using a cause-specific hazard formulation.

Further analyses are reported in the Supplement: re-estimating every summary representation on the full longitudinal record and evaluating it at the same landmark (Supplementary Table S16); complete-case and earlier-cutoff analyses (Supplementary Tables S20 and S21); refitting under four input sets, from demographics alone to all inputs, together with an eight-variable score distilled from the sequence model (Supplementary Table S10) and the share of events captured at a top-decile screening capacity (Supplementary Table S12); temporal validation trained on patients indexed before 2018 and tested on those indexed later (Supplementary Table S7); subgroup and fairness audits (Supplementary Tables S8 and S9); a learning-curve analysis (Supplementary Figure S3); a group-based trajectory model entered as a categorical covariate (Supplementary Table S22); a gap-adjusted alternative definition of average adherence (Supplementary Table S11); re-specification of the regimen-change feature block and grouping of intercorrelated features (Supplementary Tables S17 and S13); descriptive statistics for every feature (Supplementary Table S15); and an examination of whether the attention weights offer a faithful explanation[36] (Supplementary Note 1).

### Anchoring the ladder on the PREVENT equation

We repeated the ladder on top of the American Heart Association PREVENT equation, computing the published 10-year atherosclerotic cardiovascular disease score for each patient[2], recalibrating it to the three-year outcome, and adding in turn average adherence, the full engineered summary, and the fill-sequence model’s score, with folds, imputations, landmark, horizon, and bootstrap identical to the primary analysis.

## Results

### Study cohort

Of 45,017 source patients, 13,796 filled an antihypertensive prescription. Of these patients, 12,473 filled at least two prescriptions; 8,836 met the index-encounter and age criteria. In total, 7,625 patients met all eligibility criteria and formed the analytic cohort (Supplementary Figure S4). Over a median follow-up of 6.8 years, 2,626 patients (34.4%) experienced MACE (myocardial infarction, stroke, or death from any cause). Patients who experienced MACE were older (mean 62 versus 55 years), more often had prior coronary disease (22% versus 7%), and had lower kidney function (Table 1).

**Table 1:** Baseline characteristics of the study cohort.

| Characteristic | Overall<br>(N = 7,625) | No MACE<br>(n = 4,999) | MACE<br>(n = 2,626) | Missing, n (%) | SMD |
| --- | --- | --- | --- | --- | --- |
| *Demographics and clinical* |  |  |  |  |  |
| Age, years | 57.4 (12.8) | 54.9 (12.4) | 62.1 (12.2) | 0 | −0.587 |
| Systolic blood pressure, mmHg | 131.0 (19.3) | 131.4 (18.1) | 130.3 (21.2) | 3,128 (41.0) | 0.054 |
| Body-mass index, kg/m<sup>2</sup> | 31.9 (7.5) | 32.3 (7.5) | 31.2 (7.5) | 2,267 (29.7) | 0.135 |
| Total cholesterol, mg/dL | 178.7 (43.1) | 183.1 (42.2) | 169.9 (43.4) | 1,311 (17.2) | 0.308 |
| HDL cholesterol, mg/dL | 51.0 (16.6) | 52.5 (16.6) | 48.2 (16.4) | 1,322 (17.3) | 0.260 |
| eGFR (CKD-EPI 2021), mL/min/1.73 m<sup>2</sup> | 81.6 (23.9) | 86.1 (21.3) | 73.2 (26.1) | 1,033 (13.5) | 0.541 |
| Female, n (%) | 3,719 (48.8) | 2,646 (52.9) | 1,073 (40.9) | 0 |  |
| *Race, n (%)* |  |  |  | 0 |  |
| White | 6,408 (84.0) | 4,115 (82.3) | 2,293 (87.3) |  |  |
| Black or African American | 782 (10.3) | 549 (11.0) | 233 (8.9) |  |  |
| Other or unknown | 435 (5.7) | 335 (6.7) | 100 (3.8) |  |  |
| Current smoker, n (%) | 351 (4.6) | 201 (4.0) | 150 (5.7) | 0 |  |
| Smoking status unknown, n (%) | 3,551 (46.6) | 2,287 (45.7) | 1,264 (48.1) |  |  |
| *Comorbidities, treatment, and healthcare use, n (%)* |  |  |  |  |  |
| Diabetes | 1,637 (21.5) | 995 (19.9) | 642 (24.4) | 0 | −0.109 |
| Chronic kidney disease | 696 (9.1) | 319 (6.4) | 377 (14.4) | 0 | −0.262 |
| Coronary artery disease | 919 (12.1) | 332 (6.6) | 587 (22.4) | 0 | −0.446 |
| Stroke | 350 (4.6) | 63 (1.3) | 287 (10.9) | 0 | −0.404 |
| Heart failure | 512 (6.7) | 191 (3.8) | 321 (12.2) | 0 | −0.309 |
| Atrial fibrillation | 482 (6.3) | 234 (4.7) | 248 (9.4) | 0 | −0.186 |
| Statin in prior 12 months | 2,927 (38.4) | 1,762 (35.2) | 1,165 (44.4) | 0 | −0.186 |
| Encounters in prior 365 days, median [IQR] | 4 [2, 9] | 4 [2, 7] | 5 [2, 15] | 0 | −0.258 |
| *Outcomes during follow-up, n (%)* |  |  |  |  |  |
| MACE (myocardial infarction, stroke, or death from any cause) | 2,626 (34.4) | 0 (0.0) | 2,626 (100.0) |  |  |
| Myocardial infarction | 995 (13.0) | 0 (0.0) | 995 (37.9) |  |  |
| Stroke | 809 (10.6) | 0 (0.0) | 809 (30.8) |  |  |
| All-cause death | 1,641 (21.5) | 0 (0.0) | 1,641 (62.5) |  |  |

### Representations that preserve more of the medication timeline progressively improve prediction

The ladder and the prediction framework are set out in Figure 1. We quantified prognostic information lost when a patient’s antihypertensive history is collapsed into practice-based summaries. Discrimination rose monotonically as each representation preserved more of the medication timeline. Average adherence (PDC) added almost nothing to the clinical-only reference (ΔC 0.0017, 95% CI 0.0002 to 0.0032; C 0.7113 to 0.7131). Across four definitions of the outcome, including one restricted to cardiovascular causes and one excluding death entirely, this increment ranged from 0.0013 to 0.0017, against 0.0283 for the ordered fill sequence (Supplementary Table S23). Adding regimen change, then the full engineered summary, and then the ordered fill sequence produced successively larger gains (C 0.7246, 0.7273, and 0.7396). Group-based trajectory modeling, which assigns each patient to one of six discrete twelve-month coverage patterns, added 0.0004 (95% CI −0.0018 to 0.0027) over the clinical-only reference and did not exceed average adherence (Supplementary Table S22). The fill-sequence model exceeded the full engineered summary by 0.0123 (95% CI 0.0040 to 0.0202) on the primary landmark metric and by 0.0130 (95% CI 0.0075 to 0.0187) on the prespecified secondary measure, the landmark-anchored form of Antolini’s time-dependent concordance (Supplementary Table S2). We then separated the effect of the representation from that of the model class in a factorial comparison. Enriching the representation increased discrimination by 0.0181 to 0.0191 across the linear and nonlinear models. This was approximately three times the 0.0059 to 0.0078 gain from substituting the nonlinear gradient-boosted model for the linear model at a fixed representation (Supplementary Table S3). A nonlinear model on the engineered summaries produced lower discrimination than the sequence model (0.7362 versus 0.7396), and removing fill order from the sequence model lowered C to 0.7284, erasing its advantage over the engineered summary, which reaches 0.7273 on the same cohort (Supplementary Table S3).

**Figure 1:**
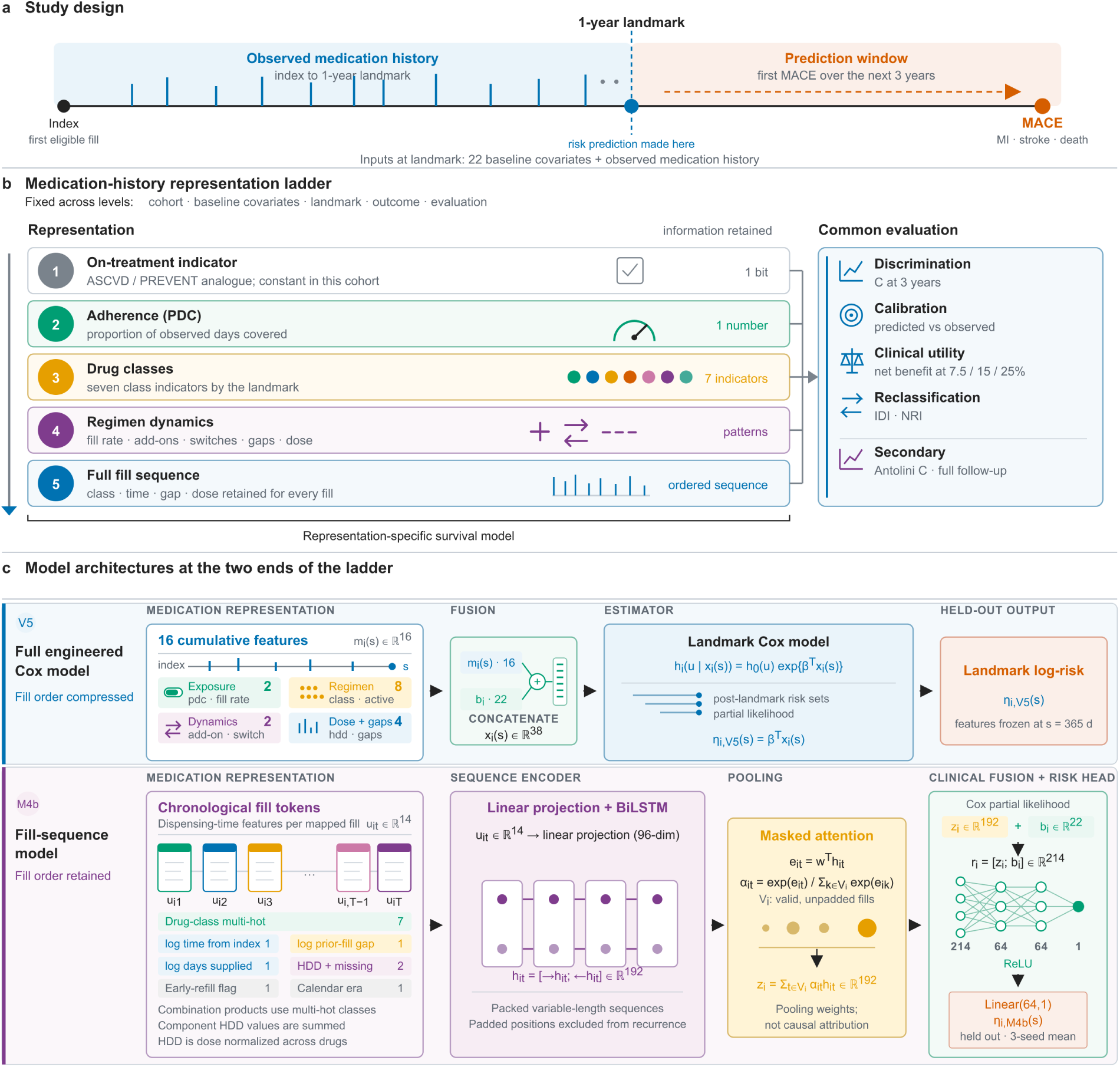
Study design and the representation ladder. (a) Single-patient prediction framework. Patients entered at their first eligible antihypertensive fill. Medication history observed from the index date to a one-year landmark was used to predict the first major adverse cardiovascular event (myocardial infarction, stroke, or death from any cause) over the following three years. Baseline clinical covariates were measured at index. (b) The representation ladder. The same fill history is summarized with increasing longitudinal detail: a single on-treatment indicator, which is constant within this cohort; average adherence; drug-class indicators at the landmark; regimen-dynamics features; and the complete ordered fill sequence. Cohort, baseline covariates, outcome, prediction window, and evaluation are held constant, and only the medication-history representation differs. (c) The two ends of the ladder. The full engineered summary compresses the record into 16 cumulative features evaluated at the landmark, whereas the fill-sequence model reads one token per mapped fill through a bidirectional long short-term memory network with attention pooling, fuses the pooled representation with the same 22 baseline covariates, and is trained with the Cox partial likelihood.

### The same ordering holds when the ladder is anchored on the PREVENT equation

The reference model above is fitted to our data and is richer than any published calculator. We therefore repeated the ladder with PREVENT as the baseline (Figure 2b; full contrast matrix in Supplementary Table S19). PREVENT alone reached C 0.6512. Adding average adherence changed discrimination by 0.0025 (95% CI −0.0003 to 0.0054), an increment not distinguishable from zero. The full engineered summary increased discrimination by 0.0317 (95% CI 0.0222 to 0.0414). The fill-sequence score increased discrimination by 0.0458 (95% CI 0.0319 to 0.0589). Within a fully treated population, substituting average adherence for the constant indicator does not recover information that the indicator cannot carry. Representations preserving refill timing and regimen evolution do.

**Figure 2:**
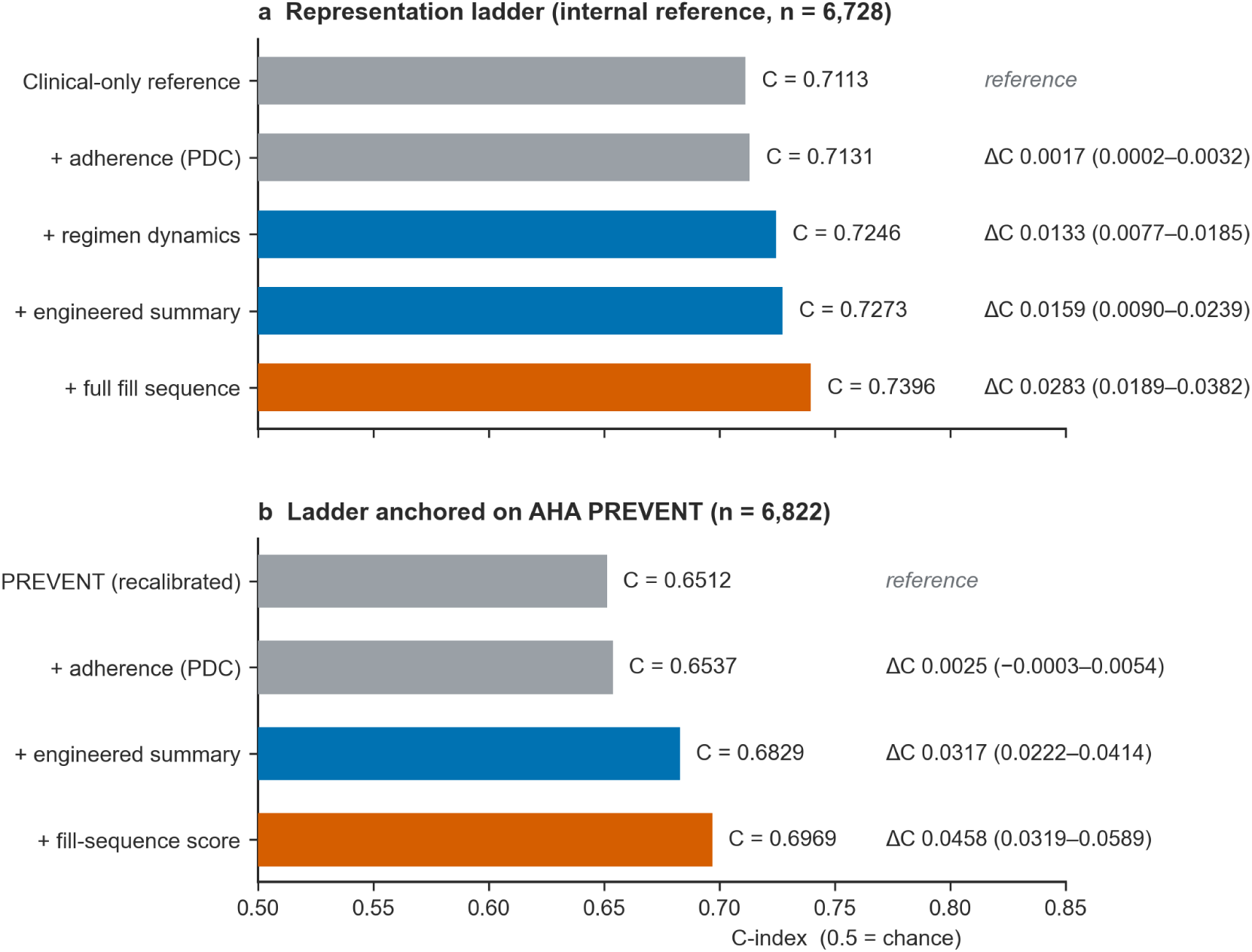
Discrimination across the representation ladder, against an internal reference and against the PREVENT equation. (a) Held-out discrimination for each representation on the intersection cohort (n = 6,728) shared by every paired comparison in this ladder. Bars are shaded by representation family: gray for the clinical-only reference and average adherence, blue for the engineered summaries, and orange-red for the fill sequence. (b) The same ladder anchored on the American Heart Association PREVENT equation, with rungs restricted to average adherence, the full engineered summary, and the fill-sequence score, in the 6,822 patients the fill-sequence model could score. Both panels plot paired-bootstrap estimates with 95% confidence intervals and begin at 0.5, the value expected by chance. The reference in (a) is fitted to this cohort and contains covariates that PREVENT does not, so it is the stronger starting point; the ordering of the rungs rather than the size of the increments is what is comparable between the panels. The nested cohorts are set out in Supplementary Methods.

### The gain is not an artifact of model complexity or fill volume

The final rung is a neural sequence model, so we examined whether its advantage reflected model flexibility or dispensing volume rather than recovered information. Model complexity was excluded by the factorial result above and by the order ablation. Nonlinearity contributed little, and discarding fill order erased the gain. The baseline covariates already included prior-year encounters, and the engineered summary already included fill rate, distinct-class count, and regimen-change counts, so the sequence-versus-summary comparison was already volume-adjusted; a volume-only model reached only C 0.7174, which the sequence model exceeded by 0.0217 (95% CI 0.0127 to 0.0310) (Supplementary Table S4). Re-estimating the summaries instead on the full longitudinal record, which is the specification more favorable to them, narrowed the difference to 0.0080 (95% CI −0.0003 to 0.0157), an interval that includes zero although the ordering is unchanged, and the advantage that specification conferred on the summaries increased with the amount of time-varying medication information each contained while leaving the medication-free reference unchanged. Because that reference carries no medication features and therefore no time-varying content, it serves as a negative control: the absence of any advantage for it identifies the effect as arising from the medication features rather than from the estimation procedure in general (Supplementary Table S16).

### Average adherence captures only part of the fill trajectory

Among patients adherent by the Medicare Star Ratings threshold (PDC ≥0.80; n = 3,240, mean PDC 0.95), PDC no longer ranked risk (within-group C 0.4628, below the 0.5 chance level). The sequence model, however, retained strong discrimination in this same band (C 0.7455, 95% CI 0.7220 to 0.7694). Because the sequence model also carries the clinical covariates, we scored three design-matched reference models on the same patients to separate the sources of that discrimination (Figure 3a; Supplementary Table S18). The clinical covariates alone reached C 0.7227 (95% CI 0.6977 to 0.7488), and adding average adherence to them left discrimination essentially unchanged, at C 0.7222, a covariate-matched restatement of the univariate result. A model given the fill sequence and no covariates at all reached C 0.6499 (95% CI 0.6211 to 0.6762), well above chance, while the full model exceeded the clinical covariates by 0.0228. The same null holds in the low-adherence band, where average adherence spans a wide range of coverage (mean 0.26), so the within-band result is not an artifact of restricted variation: average adherence discriminates between bands (whole-cohort C 0.5310) but not within them. Observed three-year MACE across model-defined risk thirds rose from 3.6% to 6.8% to 22.8% within this apparently uniform group (Figure 3b).

**Figure 3:**
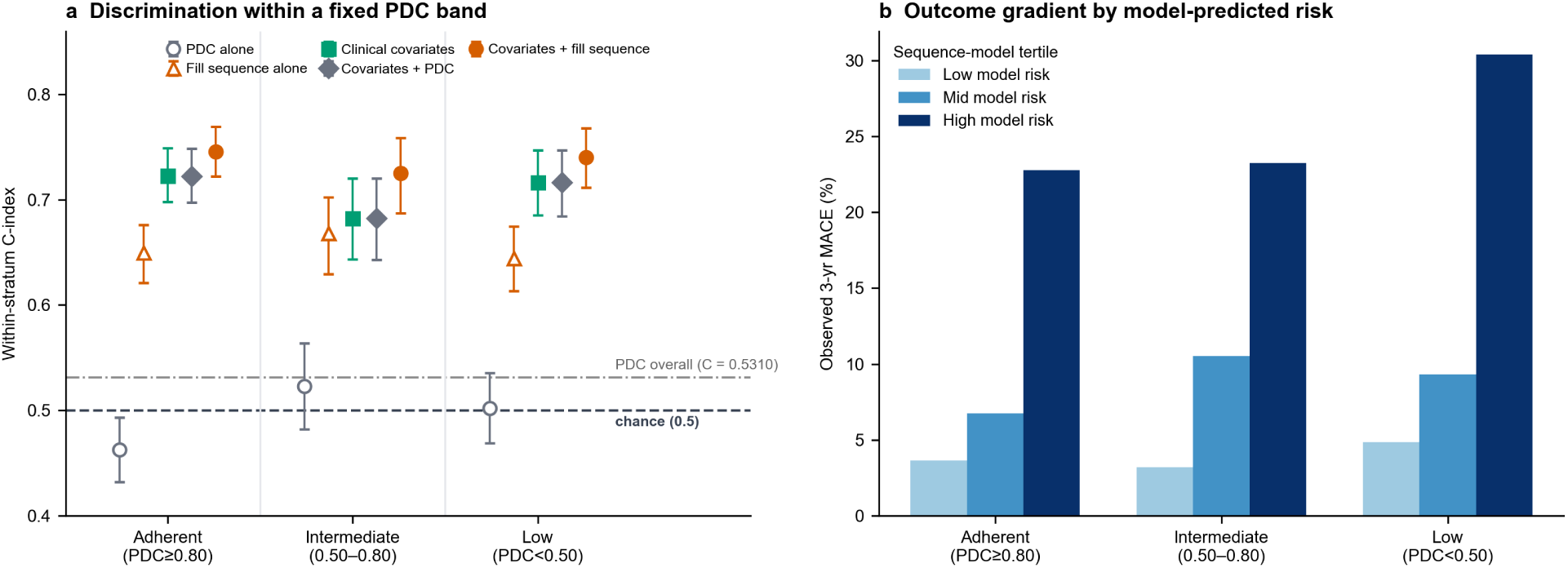
Cardiovascular risk within fixed bands of average adherence. (a) Within-band discrimination of five risk scores, all evaluated on the same patients. Average adherence (proportion of days covered, PDC) enters as a single variable, as it is used in quality measurement; the clinical covariate model and the fill-sequence model are the same models reported elsewhere in this article, and the sequence-only model uses the fill history with no covariates. Among patients classified as adherent by the Medicare Star Ratings threshold (PDC ≥0.80), average adherence did not rank three-year risk: its within-band concordance falls below the chance level of 0.5, which is marked, as is the overall concordance of PDC in the whole cohort. Markers are point estimates and bars are 95% confidence intervals. (b) Observed three-year incidence of major adverse cardiovascular events across thirds of fill-sequence model predicted risk within each band.

### Calibration and clinical utility

Calibration and decision-analytic net benefit are shown in Figure 4, with the full decile calibration and the decision-curve range in Supplementary Figures S1 and S2. Before recalibration, the fill-sequence model overpredicted at the high-risk extreme, by more than any other representation (predicted 53% versus observed 39% MACE in the highest-risk decile; calibration slope 0.905). A one-parameter recalibration estimated on held-out folds corrected this (slope 0.991) without altering patient ranking. On decision-curve analysis, the fill-sequence model provided the greatest net benefit at the screening thresholds most relevant to primary prevention (net benefit 0.061 at a 7.5% threshold). It was comparable to the engineered summary at 15%, and was lower at the 25% high-risk threshold, where its wider probability spread generated more false positives. Isotonic recalibration removed this disadvantage (Supplementary Table S6).

**Figure 4:**
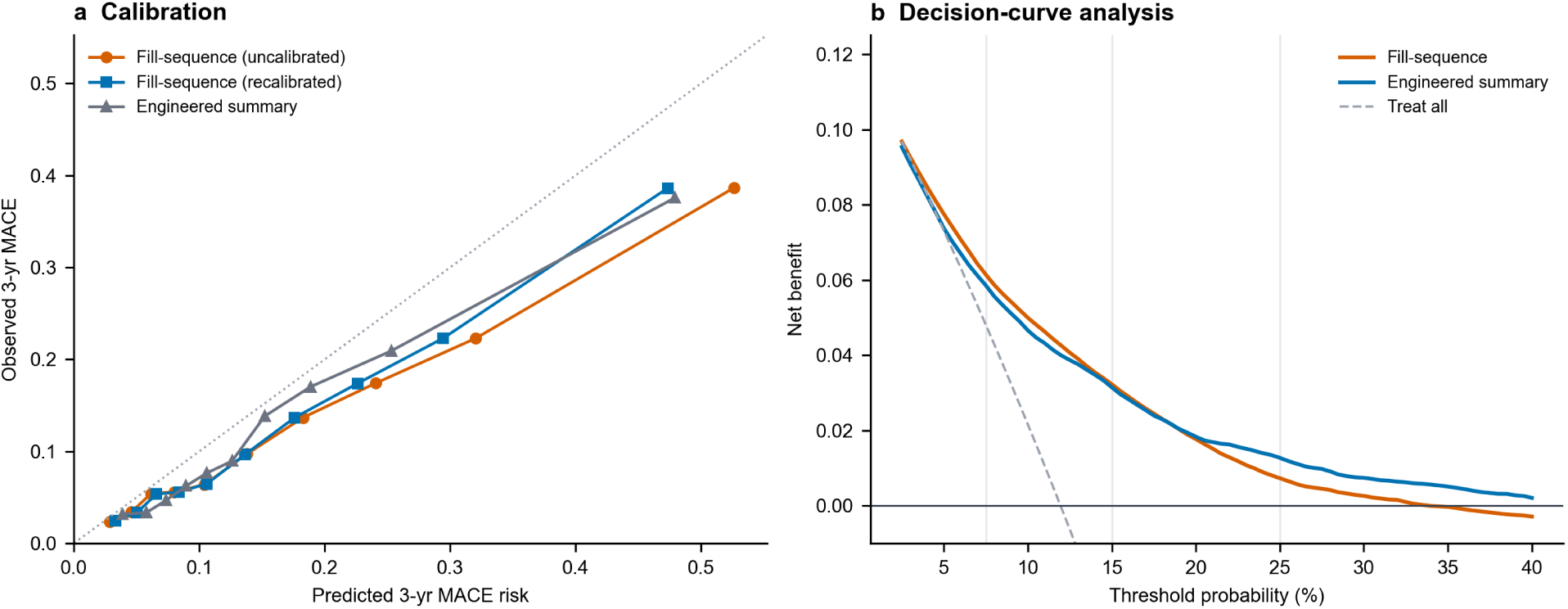
Calibration and clinical utility. (a) Calibration. Each marker is one decile of predicted risk, plotting the observed three-year incidence estimated by the Kaplan-Meier method against the mean predicted three-year risk in that decile. The dotted diagonal denotes perfect calibration; markers below it mean predicted risk exceeded observed risk. All three models follow the diagonal through the lower deciles and fall below it as predicted risk rises, so overprediction is concentrated in the highest-risk patients. A one-parameter recalibration estimated on held-out folds brings the fill-sequence calibration slope from 0.905 to 0.991 without changing ranking or discrimination. (b) Decision-curve analysis. Net benefit, expressed in true positives per patient, is plotted against threshold probability, with vertical guides at the 7.5%, 15%, and 25% thresholds. Higher curves are better, and a model is worth using at a given threshold only if it lies above both treating every patient (dashed) and treating none (the line at zero). Values are pooled across the 20 imputed datasets.

### Risk stratification when laboratory data are unavailable

Discrimination rose across four independent input sets (Figure 5): 0.6210 for demographics alone, 0.6876 for demographics with pharmacy fill history, 0.7099 for demographics with laboratory and clinical values, and 0.7391 for all inputs together (Supplementary Table S14). Pharmacy dispensing data therefore complement rather than substitute for laboratory-based assessment. A parsimonious eight-variable score distilled from the fill-sequence model, six standard clinical variables together with the number of drug classes and current use of a thiazide-type diuretic, reached C 0.7242, below both the full engineered summary and the fill-sequence model itself but with directly interpretable hazard ratios (Supplementary Table S10).

**Figure 5:**
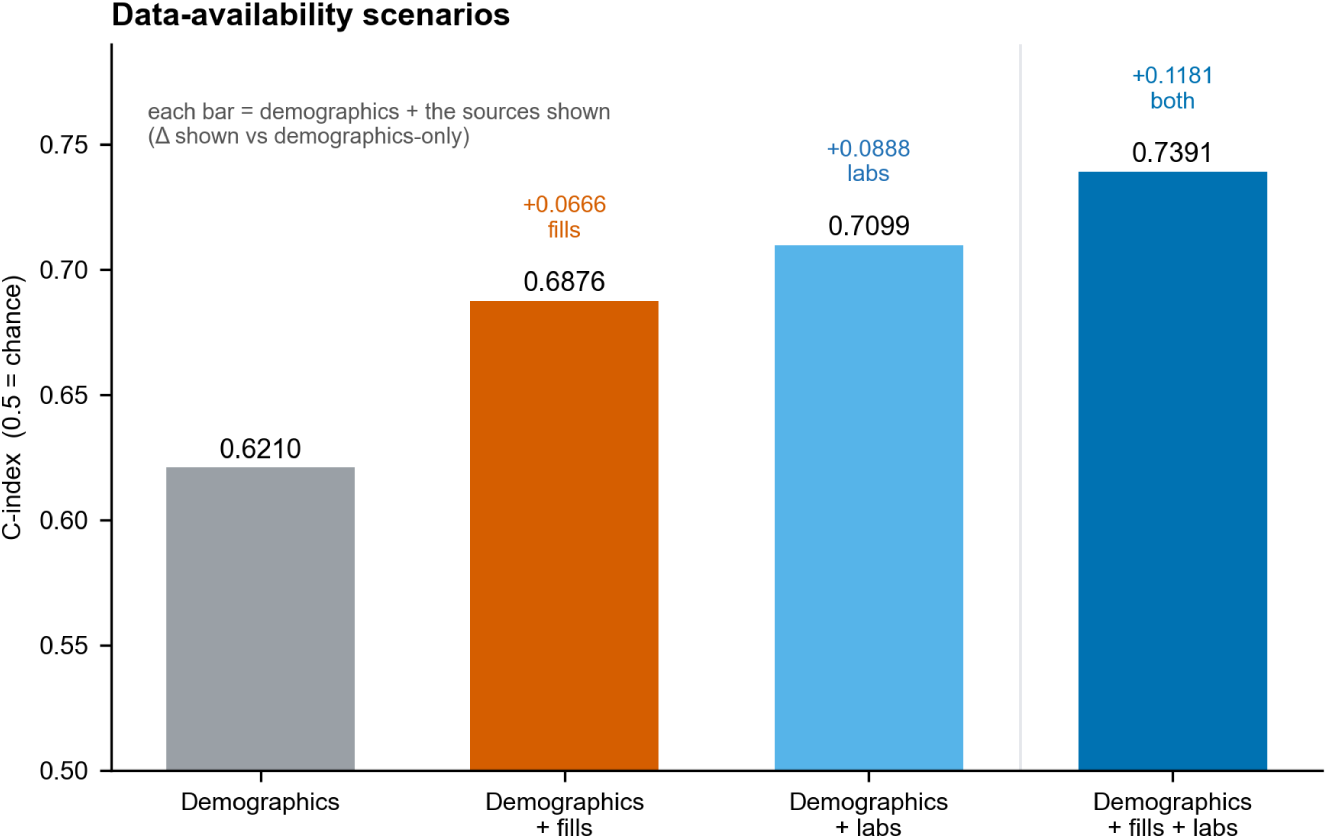
Discrimination under four data-availability scenarios. Each bar is an independent input set rather than a cumulative addition: demographics alone; demographics with pharmacy fill history; demographics with laboratory and clinical values, which is the clinical-only reference; and all inputs together. All four bars are computed on one cohort (n = 6,822) and one estimation setup. The axis begins at 0.5, the value expected by chance.

### Temporal and subgroup robustness

In temporal validation, training on the 82% of patients indexed before 2018 and testing on the 18% indexed in 2018 or later, the fill-sequence model exceeded the engineered summary by 0.0298 (95% CI 0.0077 to 0.0517), providing no evidence that the advantage weakened in the later era (Figure 6a). Exploratory sub-group analyses, not adjusted for multiplicity, localized the difference to the two lower adherence tertiles (ΔC 0.0198, 95% CI 0.0079 to 0.0320 and 0.0225, 0.0071 to 0.0373) and to single-class regimens (ΔC 0.0153, 95% CI 0.0053 to 0.0249) (Figure 6b); it was absent among the most adherent and in multi-class regimens. In the full model, discrimination was similar between men and women (C 0.7312 and 0.7362) and between White and Black patients (C 0.7351 and 0.7347, the latter with 73 events), and the richer representation moved the calibration slope among Black patients from 1.14 in the clinical-only reference to 0.98; discrimination was lower at age 65 or older than below 65 (0.6779 versus 0.7463). Full fairness results are reported in Supplementary Table S9.

**Figure 6:**
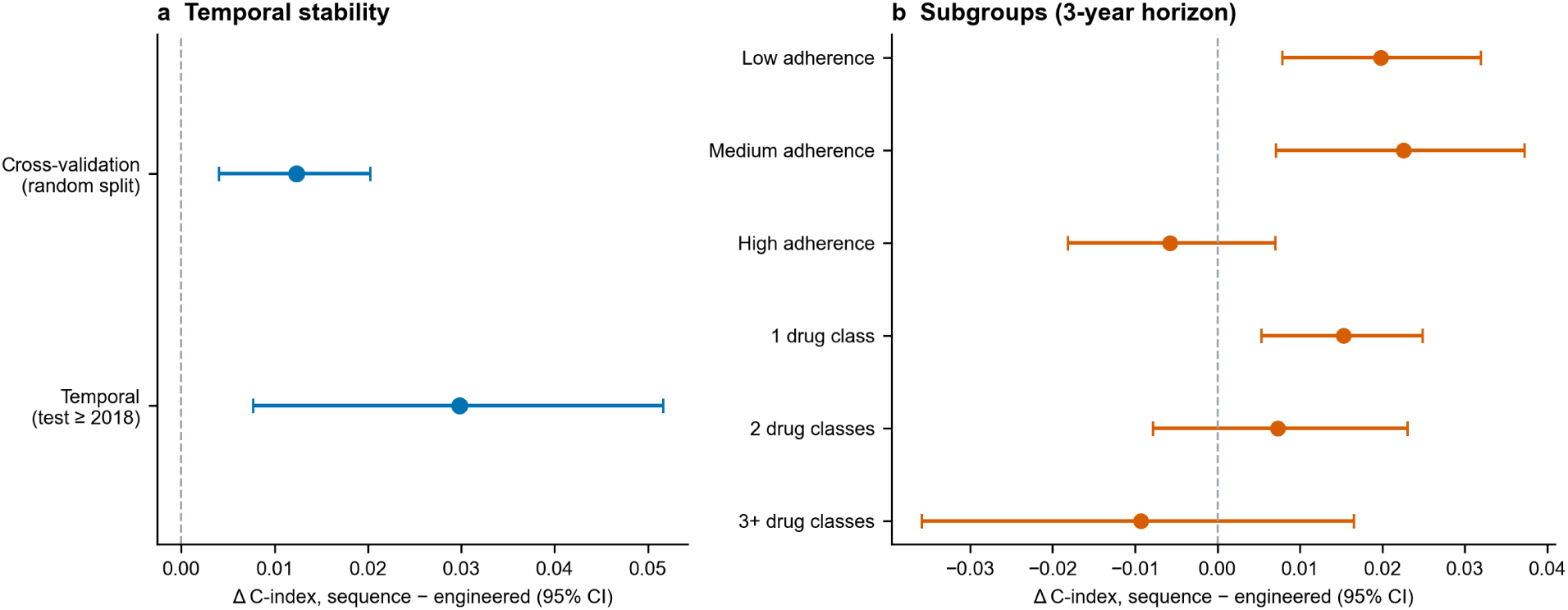
Temporal validation and subgroup differences. (a) Difference in discrimination between the fill-sequence model and the full engineered summary under random-split cross-validation and under temporal validation restricted to patients first treated in 2018 or later. (b) The same difference within subgroups defined by adherence tertile and by the number of distinct drug classes, at the three-year horizon. Both panels plot a difference and are therefore centered on zero; positive values favor the fill-sequence model. Bars are 95% confidence intervals from the paired bootstrap.

## Discussion

The central finding of this study is that the prognostic value of antihypertensive dispensing history depended strongly on how that history was represented. Average adherence added little discrimination beyond standard clinical factors and provided no meaningful risk ranking among patients classified as adherent. In contrast, progressively preserving refill timing, regularity, and regimen evolution produced progressively greater discrimination, with the ordered fill sequence performing best. Compared with the best engineered summary, the fill-sequence model improved discrimination by 0.0123 (95% CI, 0.0040–0.0202) on the primary measure and by 0.0130 (95% CI, 0.0075–0.0187) on the prespecified secondary measure. These increments are notable in the context of cardiovascular risk prediction: in the Emerging Risk Factors Collaboration, adding C-reactive protein or high-density lipoprotein cholesterol to conventional risk factors increased discrimination by 0.0039 and 0.0050, respectively[37], whereas the information examined here comes from dispensing records that are already routinely collected. The principal contribution, however, is not the superiority of a particular algorithm but the controlled representation hierarchy. By holding the cohort, outcome, clinical covariates, prediction horizon, and validation framework constant, the analysis quantifies how much prognostic information is lost as longitudinal dispensing histories are compressed into conventional summary measures.

A measurement critique of this kind invites an obvious question: if the claim is that a widely used quality metric is prognostically blind, why is a sequence model needed at all? Showing that average adherence fails to rank risk within its own adherent band requires only that metric. Showing that the information it discards remains recoverable from the same record requires a model that reads the record in its original form. The fill-sequence model is used here as an existence proof. That distinction is also why the factorial and order-ablation analyses matter: they establish that what is recovered is temporal structure in the dispensing record, not the flexibility of a more expressive model.

One alternative explanation is that the dispensing sequence detects frailty or disengagement from care rather than treatment itself. Restricting the outcome to cardiovascular events increased the advantage of the dispensing sequence when the held-out predictions were re-scored against the narrower outcome, and left it unchanged when every representation was refitted to it (Supplementary Tables S23 and S24). The intervals overlap heavily, so this is not a demonstrated difference, but it is the opposite of the direction an explanation resting on frailty or disengagement from care would predict. The finding illustrates a broader measurement principle. Two patients can have the same average value but follow markedly different trajectories, and those trajectories may contain prognostic information that the average obscures. Variation in blood pressure between visits provides information beyond mean blood pressure[38,39] and time in range captures glycemic dynamics not represented by average glycemia[40,41].

These findings clarify how PDC should be interpreted. PDC remains central to medication-quality measurement, but in this study it added little cardiovascular risk-ranking information, particularly within the adherent band where observed three-year risk varied more than sixfold. A broader lesson is that widely used metrics can conceal clinically important subgroup variation[42]. Medication-quality measurement and cardiovascular risk stratification are distinct tasks. PDC remains appropriate for the former; timing-aware representations may complement it when the goal is the latter. This was not an artifact of restricted adherence variation within the adherent band: average adherence also failed to rank risk within the low-adherence band, where it varies widely, and adding it to the clinical covariates changed discrimination by less than 0.0006 in every band.

In this fully treated cohort, the antihypertensive indicator in PREVENT is constant, and replacing it with average adherence added nothing beyond the calculator. Replacing it with representations that preserve refill timing and regimen evolution added 0.0317 to 0.0458 in discrimination. If medication information is to improve a guideline calculator in a treated population, the useful quantity appears to be the structure of the dispensing record rather than its average level.

The fill-sequence model’s advantage was concentrated among patients with lower or more irregular refill coverage, where average PDC is most likely to compress heterogeneous trajectories, and its advantage over the engineered summary was absent in the highest adherence tertile.

Discrimination was nearly identical between White and Black patients (0.7351 versus 0.7347), and the fill representation improved calibration among Black patients rather than degrading it. With 73 events in that group, however, these estimates are descriptive rather than confirmatory, and a non-significant difference is not evidence of equivalent performance. The clearer gradient was by age: discrimination was materially lower at 65 or older than below 65, which is where a deployed score would most need scrutiny.

This study has several limitations. First, it was conducted at a single academic health system, so the central measurement finding requires external validation: whether average adherence is similarly uninformative for risk ranking within conventional adherence strata, and whether longitudinal dispensing representations recover comparable prognostic information in other populations. Second, the engineered-feature and fill-sequence approaches differ in both representation and model class. Although the factorial comparisons and order-ablation analyses support temporal representation, rather than model flexibility alone, as the primary source of improvement, these analyses cannot eliminate all differences attributable to model specification. Third, the paired primary comparisons required an intersection cohort that excluded 190 of 6,918 eligible patients, including 45 of 817 events. Most of these excluded events arose from patients ineligible for the fixed 180-day comparator, creating selective removal of a small high-risk subgroup. The paired design preserves internal comparability across representations, but absolute discrimination may therefore differ in the full landmark cohort. Finally, approximately half of patients had at least one missing baseline covariate. Multiple imputation relied on an unverifiable missing-at-random assumption, although missingness patterns were consistent with differences in healthcare utilization. Importantly, complete-case analyses in 3,462 patients reproduced the near-null incremental discrimination of average adherence, indicating that this principal finding was not dependent on the imputation model.

## Conclusion

Cardiovascular risk tools and pharmacy quality measures compress antihypertensive medication history into simple summaries, discarding information about the timing, regularity, and evolution of dispensing. The consequences of this compression were clearest among patients classified as adherent by average PDC, where PDC provided little risk discrimination but richer representations of the same dispensing history retained prognostic information. Preserving more of the temporal structure of routinely collected dispensing data improved cardiovascular risk stratification beyond average adherence. Priorities for translation include independent multi-site validation, prospective evaluation of clinical and population-health impact, and assessment of how fill-history representations can complement existing cardiovascular risk models and population-health workflows.

## Author contributions

Y.Z.: conceptualization, methodology, software, formal analysis, data curation, visualization, funding acquisition, and writing (original draft). C.A.L.: conceptualization, methodology, supervision, and writing (review and editing). K.B.F., A.B.C., M.P.D., and K.N.: conceptualization, methodology, and writing (review and editing). All authors reviewed and approved the final manuscript. Roles follow the CRediT taxonomy.

## Data Availability

The electronic health record and pharmacy dispensing data analyzed in this study are not publicly available because they contain protected health information and their use is governed by the University of Michigan Medical School Institutional Review Board approval (HUM00244597) and by the agreements under which they were obtained. De-identified analytic datasets are available to only qualified researchers from the corresponding author on reasonable request, subject to institutional approval and a data-use agreement.

## Acknowledgments

Large language models were used at several stages of this work: for language editing; for drafting and revising passages of the manuscript; for checking the consistency of numerical values across the manuscript, the Supplementary Information, and the underlying analysis outputs; and for writing and debugging parts of the analysis code. All study design decisions, model specifications, and interpretations are the authors’ own, and every passage and every line of code produced with such assistance was reviewed and verified by the authors, who take full responsibility for the content of this article.

## Data availability

The electronic health record and pharmacy dispensing data analyzed in this study are not publicly available because they contain protected health information and their use is governed by the University of Michigan Medical School Institutional Review Board approval (HUM00244597) and by the agreements under which they were obtained. De-identified analytic datasets are available to qualified researchers from the corresponding author on reasonable request, subject to institutional approval and a data-use agreement.

## Code availability

The analysis code that supports the findings of this study is held in the repository github.com/franklin-yz/Dissertation_aim2_script, which is currently private. It is available from the corresponding author on reasonable request, including to editors and reviewers during peer review, and will be deposited, together with the trained weights of the fill-sequence model, in a public archive with a citable digital object identifier on acceptance.

## References

[1] Arnett DK, Blumenthal RS, Albert MA, et al. 2019 ACC/AHA guideline on the primary prevention of cardiovascular disease. Circulation. 2019;140:e596–646.

[2] Khan SS, Matsushita K, Sang Y, et al. Development and validation of the American Heart Association’s Predicting Risk of Cardiovascular Disease EVENTs (PREVENT) equations. Circulation. 2024;149:430–49.

[3] Pharmacy Quality Alliance, Centers for Medicare and Medicaid Services. Proportion of Days Covered (PDC): adherence measures for Medicare Part D Star Ratings. 2024.

[4] Peng X, Wan L, Yu B, et al. The link between adherence to antihypertensive medications and mortality rates in patients with hypertension: a systematic review and meta-analysis of cohort studies. BMC Cardiovasc Disord. Published Online First: 2025. doi: 10.1186/s12872-025-04538-6

[5] Liu M, Zheng G, Cao X, et al. Better Medications Adherence Lowers Cardiovascular Events, Stroke, and All-Cause Mortality Risk: A Dose-Response Meta-Analysis. J Cardiovasc Dev Dis. 2021;8:146.

[6] Ritchey M, Chang A, Powers C, et al. Vital signs: Disparities in antihypertensive medication nonadherence among Medicare Part D beneficiaries – United States, 2014. MMWR Morb Mortal Wkly Rep. 2016;65:967–76.

[7] Hou Q, Zhao Y, Wu Y. Medication adherence trajectories and clinical outcomes in patients with cardiovascular disease: a systematic review and meta-analysis. J Glob Health. 2025;15:04145.

[8] Librero J, Sanfélix-Gimeno G, Peiró S. Medication Adherence Patterns after Hospitalization for Coronary Heart Disease: A Population-Based Study Using Electronic Records and Group-Based Trajectory Models. PLoS One. 2016;11:e0161381.

[9] Rodríguez-Bernal CL, Sánchez-Saez F, Bejarano-Quisoboni D, et al. Assessing Concurrent Adherence to Combined Essential Medication and Clinical Outcomes in Patients With Acute Coronary Syndrome: A Population-Based, Real-World Study Using Group-Based Trajectory Models. Front Cardiovasc Med. 2022;9:863876.

[10] Wood AM, Greenland P. Evaluating the Prognostic Value of New Cardiovascular Biomarkers. Dis Markers. 2009;26:199–207.

[11] Cook NR. Use and misuse of the receiver operating characteristic curve in risk prediction. Circulation. 2007;115:928–35.

[12] Avins AL, Pressman A, Ackerson L, et al. Placebo Adherence and Its Association with Morbidity and Mortality in the Studies of Left Ventricular Dysfunction. J Gen Intern Med. 2010;25:1275–81.

[13] Murray EJ, Hernán MA. Improved adherence adjustment in the Coronary Drug Project. Trials. 2018;19:158.

[14] Coronary Drug Project Research Group. Influence of adherence to treatment and response of cholesterol on mortality in the Coronary Drug Project. N Engl J Med. 1980;303:1038–41.

[15] Surescripts, LLC. National medication-history and electronic-prescribing network. 2023.

[16] Collins GS, Moons KGM, Dhiman P, et al. TRIPOD+AI statement: updated guidance for reporting clinical prediction models that use regression or machine learning methods. BMJ. 2024;385:e078378.

[17] Inker LA, Eneanya ND, Coresh J, et al. New creatinine- and cystatin C-based equations to estimate GFR without race. N Engl J Med. 2021;385:1737–49.

[18] Charles K, Lewis MJ, Montgomery E, et al. The 2021 Chronic Kidney Disease Epidemiology Collaboration Race-Free Estimated Glomerular Filtration Rate Equations in Kidney Disease: Leading the Way in Ending Disparities. Health Equity. 2024;8:39–43.

[19] Bosco E, Hsueh L, McConeghy KW, et al. Major adverse cardiovascular event definitions used in observational analysis of administrative databases: a systematic review. BMC Med Res Methodol. 2021;21:241.

[20] Hochreiter S, Schmidhuber J. Long short-term memory. Neural Comput. 1997;9:1735–80.

[21] Bahdanau D, Cho K, Bengio Y. Neural machine translation by jointly learning to align and translate. 3rd International Conference on Learning Representations (ICLR). 2015.

[22] Choi E, Schuetz A, Stewart WF, et al. Using recurrent neural network models for early detection of heart failure onset. J Am Med Inform Assoc. 2017;24:361–70.

[23] Efron B. The efficiency of Cox’s likelihood function for censored data. J Am Stat Assoc. 1977;72:557.

[24] Austin PC, White IR, Lee DS, et al. Missing Data in Clinical Research: A Tutorial on Multiple Imputation. Can J Cardiol. 2021;37:1322–31.

[25] White IR, Royston P, Wood AM. Multiple imputation using chained equations: issues and guidance for practice. Stat Med. 2011;30:377–99.

[26] Rubin DB. Multiple Imputation for Nonresponse in Surveys. New Delhi, India: Wiley 1987.

[27] Cox DR. Regression models and life-tables. J R Stat Soc Series B Stat Methodol. 1972;34:187–202.

[28] van Houwelingen HC, Putter H. Dynamic predicting by landmarking as an alternative for multi-state modeling: an application to acute lymphoid leukemia data. Lifetime Data Anal. 2008;14:447–63.

[29] Park SY, Park JE, Kim H, et al. Review of Statistical Methods for Evaluating the Performance of Survival or Other Time-to-Event Prediction Models (from Conventional to Deep Learning Approaches). Korean Journal of Radiology. 2021;22:1697–707.

[30] Harrell FE, Lee KL, Mark DB. Multivariable prognostic models: issues in developing models, evaluating assumptions and adequacy, and measuring and reducing errors. Stat Med. 1996;15:361–87.

[31] Vickers AJ, Van Calster B, Steyerberg EW. Net benefit approaches to the evaluation of prediction models, molecular markers, and diagnostic tests. BMJ. 2016;352:i6.

[32] Antolini L, Boracchi P, Biganzoli E. A time-dependent discrimination index for survival data. Stat Med. 2005;24:3927–44.

[33] Platt J. Probabilistic outputs for support vector machines and comparisons to regularized likelihood methods. Advances in Large Margin Classifiers. MIT Press 1999:61–74.

[34] Niculescu-Mizil A, Caruana R. Predicting good probabilities with supervised learning. Proceedings of the 22nd International Conference on Machine Learning (ICML). 2005:625–32.

[35] Chen T, Guestrin C. XGBoost: a scalable tree boosting system. Proceedings of the 22nd ACM SIGKDD International Conference on Knowledge Discovery and Data Mining. 2016:785–94.

[36] Jain S, Wallace BC. Attention is not Explanation. Proceedings of NAACL-HLT 2019. 2019:3543–56.

[37] Emerging Risk Factors Collaboration, Kaptoge S, Di Angelantonio E, et al. C-reactive protein, fibrinogen, and cardiovascular disease prediction. N Engl J Med. 2012;367:1310–20.

[38] Stevens SL, Wood S, Koshiaris C, et al. Blood pressure variability and cardiovascular disease: systematic review and meta-analysis. BMJ. 2016;354:i4098.

[39] Rothwell PM, Howard SC, Dolan E, et al. Prognostic significance of visit-to-visit variability, maximum systolic blood pressure, and episodic hypertension. Lancet. 2010;375:895–905.

[40] Yoo JH, Kim JH. Time in Range from Continuous Glucose Monitoring: A Novel Metric for Glycemic Control. Diabetes Metab J. 2020;44:828–39.

[41] Battelino T, Danne T, Bergenstal RM, et al. Clinical targets for continuous glucose monitoring data interpretation: recommendations from the international consensus on time in range. Diabetes Care. 2019;42:1593–603.

[42] Obermeyer Z, Powers B, Vogeli C, et al. Dissecting racial bias in an algorithm used to manage the health of populations. Science. 2019;366:447–53.

