## Supplementary Materials for "Digital Treatment Signatures: Cardiovascular Risk Prediction Using Antihypertensive Medication Fill History"

---

#### General notes

All reported values are held-out (cross-validated) predictions unless stated otherwise. Discrimination is the dynamic-landmark concordance at landmark s = 365 days and horizon +3 years unless another landmark or horizon is named. Paired model comparisons are computed on the intersection cohort of patients with valid predictions from every compared model; the applicable denominator is stated in each table legend. This intersection (n = 6,728) is reached from the 6,918 patients alive at the day-365 landmark by removing two groups of about 95 patients each, for two different reasons; one patient falls in both groups, which is why the two reductions total 190 rather than 191. The first is absent from the fixed 180-day landmark model, which is defined on a differently constructed eligible population. The second cannot be scored by the fill-sequence model, which requires at least one fill in the first year that maps to one of the seven modeled drug classes; a small number of patients have no such fill and therefore no sequence to read, whereas the Cox models still contribute a prediction for them from the covariates with medication features set to zero. Requiring a valid held-out prediction from every compared model therefore leaves 6,728 patients, and the drop reflects model eligibility rather than any loss of data. Absolute concordance differs by at most 0.0030 between the full-cohort and intersection-cohort denominators, and this does not affect any ranking. Concordance and differences in concordance are reported to four decimal places throughout, except where a table legend states otherwise. Interval estimates are 95% percentile bootstrap intervals from B = 1000 paired resamples of patients. A difference in concordance is the mean of the resample-level differences and is rounded independently of the concordances it is derived from, so a reported difference may depart by 0.0001 from the difference of two rounded endpoints.

Model shorthand: **L0**, clinical-only reference; **V1**, average adherence (PDC); **V2**, drug-class indicators at the landmark; **V3**, drug-class indicators at baseline; **V4**, adherence and regimen change; **V5**, full engineered summary; **M4b**, fill-sequence model (bidirectional LSTM with attention pooling); **M1**, fixed 180-day landmark Cox model. The M and V prefixes are retained from the analysis code and are not sequential.

**Estimation setup.** The primary analysis estimates every summary representation as an ordinary Cox model on the day-365 feature snapshot against the post-landmark outcome, which is the task the fill-sequence model is trained on, so that the comparison isolates the representation of medication history rather than the estimation setup. Unless a table states otherwise, all summary-model values in these materials are from that landmark estimation. The alternative counting-process estimation on the full longitudinal record is reported as a sensitivity analysis in Supplementary Table S16.

Supplementary Table S14. Deployment decomposition, snapshot baseline, and external benchmark 27

#### Abbreviations

ACE, angiotensin-converting enzyme; AHA, American Heart Association; ASCVD, atherosclerotic cardiovascular disease; BP, blood pressure; BIC, Bayesian information criterion; C, concordance index; CI, confidence interval; CKD, chronic kidney disease; CKD-EPI, Chronic Kidney Disease Epidemiology Collaboration; eGFR, estimated glomerular filtration rate; GBM, gradient-boosting machine; GBTM, group-based trajectory model; HbA1c, glycated hemoglobin; HDD, hypertension daily dose (dose normalized to the maximum beneficial dose); HDL, high-density lipoprotein; IBS, integrated Brier score; IDI, integrated discrimination improvement; IQR, interquartile range; LOFO, leave-one-fold-out; LSTM, long short-term memory; MACE, major adverse cardiovascular event (three-point composite of first myocardial infarction, stroke, or death from any cause); MICE, multivariate imputation by chained equations; NB, net benefit; NRI, net reclassification improvement; O/E, ratio of observed to expected events; MI, myocardial infarction; PDC, proportion of days covered; PREVENT, Predicting Risk of Cardiovascular Disease EVENTs; PPV, positive predictive value; SD, standard deviation; SHAP, Shapley additive explanations; SMD, standardized mean difference; VIF, variance inflation factor.

#### Supplementary Methods

##### S-Methods 1. Cohort construction and time-varying feature engineering

Patients entered at the first antihypertensive fill preceded by a clinical encounter within the prior year. We additionally required at least two antihypertensive fills, age ≥18 years, and non-negative post-index follow-up. Time-varying medication features were built on a 30-day interval scaffold (684,697 intervals across 7,625 patients). They accumulated over the half-open window [index, t) at each interval, so that no feature used information after the prediction time. Each fill was mapped to one of seven antihypertensive classes (ACE inhibitors, angiotensin-receptor blockers, calcium-channel blockers, beta-blockers, thiazide-type diuretics, other diuretics, other antihypertensives), and combination products were decomposed to their component classes. The proportion of days covered (PDC) was the cumulative union-exposure PDC, the fraction of days in [index, t) on which any antihypertensive supply was available, with overlapping coverage counted once. A gap-adjusted alternative definition, in which a coverage gap terminates the exposure interval, yielded near-identical predictions (Supplementary Table S11). Regimen-dynamics features included cumulative new-class additions, class switches, refill rate, distinct-class count, dose intensity (hypertension daily dose, normalized to the maximum beneficial dose), and the longest coverage gap. In the primary analysis each summary representation was estimated as an ordinary Cox model on the patient's feature snapshot at the day-365 landmark against the post-landmark outcome, which is the task the fill-sequence model is trained on. Estimation on the full longitudinal record by counting-process methods[10] is reported as a sensitivity analysis (Supplementary Table S16).

##### S-Methods 2. Fill-token schema and sequence-model architecture

The fill-sequence model read an ordered sequence of per-fill tokens. The lean token (14 dimensions) contained only information available at dispensing and carried no engineered summary: drug-class multi-hot (7), log time since index, log gap since the previous fill, log days supplied, dose intensity, a dose-missing flag, an early-refill flag, and a calendar-era term. A bidirectional long short-term memory network (hidden size 96) with an attention-pooling head summarized the sequence to a fixed vector. This vector was concatenated with the 22 baseline clinical covariates and passed through a two-layer multilayer perceptron to a single risk score, trained with the Cox partial-likelihood objective using Efron's approximation for tied event times.[5] We chose a recurrent architecture rather than a transformer because the sequences are short: the median patient contributes 17 dispensing tokens (IQR 6 to 39), and self-attention offers its largest advantage over long contexts, which this task does not present. Architecture is also not the quantity this study varies. Optimization used AdamW with a cosine learning-rate schedule and early stopping. The production grid comprised 20 imputations × 5 folds × 3 random seeds (300 fits), and per-patient held-out risk scores were averaged over the three seeds. Features that duplicated the engineered summaries were deliberately excluded from the token, so that any advantage over the engineered summary reflected the ordering and timing of fills rather than additional variables.

##### S-Methods 3. Multiple imputation

Systolic blood pressure (41% missing), body-mass index (30%), and total cholesterol, HDL cholesterol, and creatinine (13% to 17%) were multiply imputed with `miceforest` (LightGBM backend), using m = 20 datasets, 5 MICE iterations, and random seed 42. Estimated glomerular filtration rate was recomputed from imputed creatinine using the 2021 CKD-EPI equation.[6] Smoking status (four levels including Unknown) and race (Unknown category) were not imputed. All analyses ran in each imputed dataset and were pooled by Rubin's rules. Because every model shares the same imputed covariates, full-cohort imputation does not bias the representation comparison. Imputation was not repeated within folds, and we report this as a limitation. Missingness was associated with observed visit frequency, a pattern consistent with the missing-at-random assumption.

##### S-Methods 4. Calibration of cross-validated linear predictors

Absolute risks were obtained by calibrating each model's cross-validated risk score to the observed survival function, a Kaplan-Meier calibration of the centered linear predictor. This addressed a known limitation of the survival-analysis software, whose counting-process baseline cumulative hazard for entry-time (left-truncated) Cox models is mis-scaled. The workaround recovers correctly scaled absolute probabilities from the correctly ranked linear predictor.

##### S-Methods 5. Recalibration

The fill-sequence model was recalibrated by leave-one-fold-out (LOFO) estimation within each imputation. For each held-out fold, a correction was fit on the other four folds and applied to the held-out fold, so no patient informed its own correction. Two corrections were compared: an overall scaling correction (a Cox slope, "Platt") and a flexible monotonic correction ("isotonic"). Both preserve patient ranking, so discrimination, concordance, and the paired bootstrap are unchanged by construction. Only calibration-scale metrics differ. The scaling-corrected model is used for all absolute-risk statements in the main text.

##### S-Methods 6. Sample size and events per variable

We did not perform an a priori sample-size calculation, and we analyzed all eligible patients. The primary landmark evaluation window (one-year landmark, three-year horizon) contained 817 events, approximately 21 per candidate predictor in the largest regression model. In the sensitivity analysis that estimates the same models on the full counting-process record, all 2,626 events contribute to model fitting, approximately 69 events per predictor. Both exceed the conventional minimum of 10 events per variable.[11,12]

##### S-Methods 7. Secondary-analysis specifications

- **Representation × algorithm factorial.** Each representation rung (L0, V1, V4, V5) was fit with both the linear Cox model and a nonlinear gradient-boosted survival model (XGBoost `survival:cox`) on the identical feature set, isolating the representation axis (down a rung) from the algorithm axis (linear vs nonlinear). An unordered "bag-of-fills" variant of the sequence model averaged the same per-fill information without order.
- **Fill-volume confound.** A volume-only model used counts of fills and encounters. A second model added the raw fill count to the full engineered summary. Both were compared with the fill-sequence model by paired bootstrap.
- **Within-PDC heterogeneity.** Patients were stratified into adherence bands (PDC ≥0.80, 0.50 to 0.80, and <0.50). Within each band we computed the concordance of PDC alone and of the fill-sequence

model, and the observed 3-year MACE incidence (Kaplan-Meier) across thirds of sequence-model predicted risk. PDC enters this comparison as a single variable, which is how it is used in quality measurement, whereas the fill-sequence model also carries the 22 baseline covariates. To make the attribution explicit we scored three design-matched reference models on the same patients within each band: the clinical covariates alone (L0), the clinical covariates with PDC added (V1), and the fill sequence with no covariates. All five scores are cross-validated out-of-fold predictions evaluated on one common cohort (Supplementary Table S18).

- **Reclassification and clinical utility.** Integrated discrimination improvement and net reclassification improvement[3] were computed relative to both the adherence-only model and the full engineered summary, and net benefit was assessed by decision-curve analysis[2] at treatment thresholds of 7.5%, 15%, and 25% (Supplementary Tables S5 and S6).
- **Deployment scenarios.** Models were refit with restricted inputs: demographics only (age, sex, race); demographics + fill history (no laboratory or blood-pressure values, a community-pharmacy scenario); demographics + laboratory/clinical values (the clinical-only reference, L0); and all inputs. A sequence-only model used the fill timeline with no clinical covariates.
- **Interpretable surrogate and deployable score.** A transparent linear model was fit to reproduce the fill-sequence model's risk score from 38 interpretable features (22 covariates + 16 engineered medication features). The out-of-fold $R^2$ quantifies the reproducible share. The eight features of the parsimonious score were not chosen by hand: starting from the same 38-feature pool, features were added one at a time by greedy forward selection, at each step retaining the single feature that most increased the surrogate's out-of-fold $R^2$ relative to the model's risk score. Selection stopped at eight, where the reproduced discrimination had plateaued; six of the eight are standard clinical risk factors and two are medication-derived. The resulting eight-feature Cox score was then fit directly to MACE for deployment (Supplementary Table S10).
- **Temporal validation.** Models were trained on patients indexed before 2018 and tested on those indexed in 2018 or later. The fill-sequence model and the engineered summary were compared by paired bootstrap restricted to the test patients common to both (n = 1,207). The cutoff was repeated at earlier, better-balanced splits (Supplementary Table S21).
- **Subgroups and fairness.** Differences in discrimination (M4b – V5) were estimated within adherence tertiles and regimen-complexity strata. Discrimination and calibration were compared across race, sex, age, and diabetes status. These subgroups use adherence tertiles, which are distinct from the fixed Star Ratings PDC bands used for the within-band analysis above.
- **Learning curve.** The fill-sequence model was refit on 12.5%, 25%, 50%, and 100% of the training data (test folds full size) to assess data adequacy.
- **Complete-case sensitivity.** The analysis was repeated using only the patients for whom every model covariate was recorded, without imputation (Supplementary Table S20).
- **Cause-specific outcome.** Using the underlying cause of death recorded in the state death index, a cardiovascular outcome was defined as the first myocardial infarction, stroke, or death with an underlying cause in ICD-10 chapter I, censoring non-cardiovascular deaths. Held-out predictions from every representation were first re-scored against that outcome without retraining (Supplementary Table S23), and every representation was then refitted against it using a cause-specific hazard formulation,

in which only the event indicator and the censoring time changed and the partial-likelihood objective was otherwise unaltered; subdistribution hazard models were not used (Supplementary Table S24). To contain computation, refitting used the first five imputed datasets and a single training run per fold, with the fold assignments of the primary analysis retained, a reduction applied identically to every representation. Because non-cardiovascular death is an informative censoring event, these analyses address the relative discrimination of the representations rather than absolute risk.

- **Group-based trajectory modeling.** A group-based trajectory model was fitted to twelve monthly PDC values spanning the index date to the landmark. Because monthly coverage is bounded and concentrated at its limits, a censored-normal specification was used, selecting the number of groups and the polynomial order by Bayesian information criterion within a prespecified range, subject to each group containing at least 5% of the cohort with a mean posterior assignment probability of at least 0.70. Models were fitted within training folds and held-out patients were assigned by posterior probability; because dispensing records were complete and not imputed, group assignments were identical across imputed datasets. Modal group membership then entered the landmark Cox model as a categorical covariate alongside the same clinical covariates used for every other representation (Supplementary Table S22).
- **Medication-change representation.** Counting class switches and add-ons requires an operational rule with several arbitrary parameters: the supply overlap that separates an add-on from a switch, the gap after which a class is treated as newly appearing, and the horizon over which a later refill is taken as evidence of continued therapy. Recounting class switches over the full record across a grid of these parameters showed the count to be highly definition-dependent (from roughly 2,500 to 52,000 events), which motivated testing whether the main results depend on any particular definition. The switch/add-on block was re-specified three ways, holding cohort, folds, covariates, horizon, and metric fixed: removed entirely; replaced by a corrected classification in which a returning class is a reinitiation and only a genuinely different class is a switch, computed from history up to the landmark only (so the cumulative count is a proper function of the past); and replaced by continuous measures of regimen concurrency that impose no threshold, namely the fraction of covered time on two or more classes, excess class-days per 30 days, and the signed supply overlap at each class entry. Each re-specification was compared with the published block by paired bootstrap. The supply-overlap threshold used by the corrected classification (30 days) is a software convention, the combination window of the OHDSI TreatmentPatterns package,[7] rather than an empirically validated value; the sensitivity grid is reported because no such validation exists (Supplementary Table S17). Because that block is defined over the whole record, the re-specification was carried out under the time-updated estimation, and every value in that table, including the fill-sequence model, is estimated the same way.
- **Time-updated estimation.** Summary models and sequence models can differ in how they are estimated as well as in how medication history is represented, and the primary analysis holds the estimation setup fixed so that only the representation varies. As a sensitivity analysis, every summary representation was re-estimated on the full longitudinal record by counting-process methods,[10] so that its coefficients describe the association between a patient's current medication status and the immediate hazard, and was then evaluated at the same landmark. Cohort, fold assignment (seed 42), 20 imputations, penalizer, and the primary metric were unchanged. Because L0 contains no medication

features and therefore no time-varying content, the L0 contrast serves as a negative control isolating the estimation effect itself (Supplementary Table S16).

- **Feature intercorrelation.** Several engineered summary features are intercorrelated, so their contributions are interpreted by conceptual group (adherence, regimen change, dose, and class composition) rather than individually (Supplementary Table S13). Descriptive statistics for every engineered feature and for the per-fill features read by the sequence model are given in Supplementary Table S15.

##### S-Methods 8. The two reference models and the PREVENT anchor

Two reference models are used in this article, and the distinction between them governs how every increment should be read. The clinical-only reference is fitted to this cohort's own outcome by the same cross-validated procedure as every rung above it. That is what makes the ladder a measurement of representation: because the reference already absorbs whatever a well-specified clinical model can extract from the same data, what remains for the medication representation to add is attributable to the representation rather than to a reference poorly matched to the task. The cost of that design is that no one uses this reference in practice; the instrument clinicians use is a published equation with fixed coefficients. We therefore repeated the ladder on top of the American Heart Association PREVENT equation,[4] with its rungs restricted to average adherence, the full engineered summary, and the fill-sequence score.

The two references bracket the quantity of interest. The internal reference is the harder baseline to improve upon and therefore yields the conservative estimate of representational value. The external one is deployment-relevant, but its increments are enlarged by differences that have nothing to do with how the medication history is represented. The two differ in three ways. First, the clinical-only reference is fitted to these data, whereas PREVENT's coefficients are external and fixed. Second, the clinical-only reference additionally contains prior cardiovascular disease, the rest of the comorbidity block, prior-year encounter count, race, and body-mass index, none of which enters PREVENT, a race-free primary-prevention equation; both contain age, sex, blood pressure, lipids, kidney function, smoking, and diabetes. Third, PREVENT was developed to predict 10-year atherosclerotic cardiovascular disease, whereas the outcome here is a three-year composite that includes all-cause death. Each difference makes PREVENT the weaker starting point in this cohort, so increments measured on top of it are expected to be larger. We therefore treat agreement in the ordering of the rungs as the robustness claim, and we do not read the size of an anchored increment as evidence about the size of the corresponding internal one. The fill-sequence score enters as a score rather than as a variable because a sequence cannot be represented as a single term in an equation. For the reasons above, this analysis compares how well each model ranks patients and not the accuracy of each model's absolute risks, and it is reported as an external anchor for the primary ladder rather than as an evaluation of PREVENT in its intended use.

The anchored analysis used the 6,822 patients whom the fill-sequence model could score at the landmark, the same cohort as the within-band analysis in Supplementary Table S18. The three landmark-stage denominators in this article are strictly nested, and the two reductions have different causes. Of the 6,918 patients alive at the landmark, 96 could not be scored by the fill-sequence model, which requires at least one fill mapping to a modeled drug class within the first year, leaving the 6,822 analyzed here. A further 94 of those 6,822 are absent from the fixed 180-day landmark model, which is defined on a differently constructed eligible population; 95 patients in all are absent from it, one of whom is already among the 96 above. That leaves the 6,728-patient intersection required when every model in a comparison must contribute a prediction. The PREVENT equation itself was computable for every patient and did not restrict the cohort. The full contrast matrix and the imputation-level spread are given in Supplementary Table S19.

**S-Methods 9. Pre-registration, reporting, and software**

The analysis plan (primary metric, paired-bootstrap unit, the prespecified secondary status of Antolini's time-dependent concordance,[8] and the interpretation matrix) was registered internally before model fitting. Reporting follows TRIPOD+AI.[9] Analyses used Python (lifelines for Cox models, PyTorch for the sequence model, XGBoost for the gradient-boosted comparator, scikit-learn for recalibration and surrogates, miceforest for imputation). Package versions are recorded in the environment specification included with the analysis code, which is available at the project repository (released on acceptance).

#### Supplementary Note 1. Attention is not a faithful explanation

We examined whether the fill-sequence model's attention weights identify which fills drive its predictions. Across every view examined (fill position, drug class, gap status, and patient archetype), attention deviated less than 5% from a uniform 1/(sequence length) baseline. A univariate predictor formed by weighting refill gaps by the model's own attention was non-discriminating (concordance 0.48, i.e., no better than chance). Attention is therefore diffuse and does not constitute a faithful explanation of the model, consistent with prior work.[1] We deliberately do not present an attention heatmap, which would be near-uniform and misleading. Interpretability instead rests on the transparent surrogate, which reproduces $R^2 = 0.822$ of the model's ranking out of fold, and on the eight-feature deployable score (Supplementary Table S10).

We favored this global surrogate over local feature-attribution methods such as SHAP for two reasons. First, the model's input is a variable-length sequence of fill tokens rather than a fixed tabular vector, so per-feature attributions would apportion importance across token positions rather than across clinician-interpretable variables, and would not answer the question of interest, which is how much of the model's ranking is reproducible from interpretable features versus irreducibly sequential; the surrogate's $R^2$ answers this directly, and an attribution method does not. Second, applied to the transparent linear surrogate itself an attribution method reduces to the standardized regression coefficients, which are already reported, so it would add no information.

#### Supplementary Tables

##### Supplementary Table S1. Full cohort characteristics

Final medication-backbone cohort (N = 7,625), observed (pre-imputation) values. This is the complete characterization; main-text Table 1 is the trimmed version restricted to modeled variables.

| Characteristic | Overall (N=7,625) | No MACE (n=4,999) | MACE (n=2,626) | Missing, n (%) | SMD |
|---|---|---|---|---|---|
| **Demographics & clinical** | | | | | |
| Age, years | 57.4 (12.8) | 54.9 (12.4) | 62.1 (12.2) | 0 | −0.587 |
| Systolic BP, mmHg | 131.0 (19.3) | 131.4 (18.1) | 130.3 (21.2) | 3,128 (41.0) | 0.054 |
| Body-mass index, kg/m² | 31.9 (7.5) | 32.3 (7.5) | 31.2 (7.5) | 2,267 (29.7) | 0.135 |
| Total cholesterol, mg/dL | 178.7 (43.1) | 183.1 (42.2) | 169.9 (43.4) | 1,311 (17.2) | 0.308 |
| HDL cholesterol, mg/dL | 51.0 (16.6) | 52.5 (16.6) | 48.2 (16.4) | 1,322 (17.3) | 0.260 |
| Creatinine, mg/dL | 1.1 (0.9) | 1.0 (0.7) | 1.3 (1.1) | 1,033 (13.5) | −0.288 |
| eGFR (CKD-EPI 2021), mL/min/1.73 m² | 81.6 (23.9) | 86.1 (21.3) | 73.2 (26.1) | 1,033 (13.5) | 0.541 |
| HbA1c, %[a] | 7.9 (1.7) | 7.9 (1.7) | 7.9 (1.7) | 6,481 (85.0) | −0.024 |
| **Sex**, n (%) | | | | 0 | |
| Female | 3,719 (48.8) | 2,646 (52.9) | 1,073 (40.9) | | |
| Male | 3,906 (51.2) | 2,353 (47.1) | 1,553 (59.1) | | |
| **Race (full categories)**, n (%) | | | | 22 (0.3) | |
| African American | 782 (10.3) | 549 (11.0) | 233 (8.9) | | |
| American Indian | 36 (0.5) | 21 (0.4) | 15 (0.6) | | |
| Asian | 177 (2.3) | 143 (2.9) | 34 (1.3) | | |
| Native Hawaiian and Other Pacific Islander | 7 (0.1) | 7 (0.1) | 0 (0.0) | | |
| White | 6,408 (84.0) | 4,115 (82.3) | 2,293 (87.3) | | |
| Other | 143 (1.9) | 107 (2.1) | 36 (1.4) | | |
| Patient Refused | 25 (0.3) | 21 (0.4) | 4 (0.2) | | |
| Unknown | 25 (0.3) | 18 (0.4) | 7 (0.3) | | |
| **Ethnicity**, n (%) | | | | 209 (2.7) | |
| Hispanic | 163 (2.1) | 120 (2.4) | 43 (1.6) | | |
| Non-Hispanic | 7,054 (92.5) | 4,562 (91.3) | 2,492 (94.9) | | |
| Patient refused | 55 (0.7) | 47 (0.9) | 8 (0.3) | | |
| Unknown | 144 (1.9) | 108 (2.2) | 36 (1.4) | | |
| **Smoking status**, n (%) | | | | 0 | |
| Current | 351 (4.6) | 201 (4.0) | 150 (5.7) | | |
| Former | 1,555 (20.4) | 916 (18.3) | 639 (24.3) | | |
| Never | 2,168 (28.4) | 1,595 (31.9) | 573 (21.8) | | |
| Unknown | 3,551 (46.6) | 2,287 (45.7) | 1,264 (48.1) | | |
| **Comorbidities & treatment**, n (%) | | | | | |
| History of hypertension | 3,298 (43.3) | 2,190 (43.8) | 1,108 (42.2) | 0 | 0.033 |
| History of diabetes | 1,637 (21.5) | 995 (19.9) | 642 (24.4) | 0 | −0.109 |
| Diabetes, composite flag[b] | 2,612 (34.3) | 1,595 (31.9) | 1,017 (38.7) | 0 | −0.143 |
| History of CKD | 696 (9.1) | 319 (6.4) | 377 (14.4) | 0 | −0.262 |
| History of coronary artery disease | 919 (12.1) | 332 (6.6) | 587 (22.4) | 0 | −0.446 |
| History of stroke | 350 (4.6) | 63 (1.3) | 287 (10.9) | 0 | −0.404 |
| History of heart failure | 512 (6.7) | 191 (3.8) | 321 (12.2) | 0 | −0.309 |
| History of atrial fibrillation | 482 (6.3) | 234 (4.7) | 248 (9.4) | 0 | −0.186 |
| Statin in prior 12 mo | 2,927 (38.4) | 1,762 (35.2) | 1,165 (44.4) | 0 | −0.186 |

*continued on next page*

| Characteristic | Overall (N=7,625) | No MACE (n=4,999) | MACE (n=2,626) | Missing, n (%) | SMD |
|---|---|---|---|---|---|
| Statin ever | 3,345 (43.9) | 2,031 (40.6) | 1,314 (50.0) | 0 | −0.189 |
| **Healthcare utilization & follow-up**, median [IQR] | | | | | |
| Encounters in prior 365 d | 4 [2, 9] | 4 [2, 7] | 5 [2, 15] | 0 | −0.258 |
| Comorbidity count | 0 [0, 1] | 0 [0, 1] | 1 [0, 2] | 0 | −0.393 |
| Follow-up, days | 2,473 [1,597, 3,606] | 2,597 [1,792, 3,720] | 2,230 [1,197, 3,349] | 0 | 0.284 |
| Time to MACE or censoring, days | 2,302 [1,270, 3,446] | 2,597 [1,792, 3,720] | 1,502 [613, 2,712] | 0 | 0.680 |
| **Outcomes during follow-up**, n (%) | | | | | |
| MACE (3-point composite) | 2,626 (34.4) | 0 (0.0) | 2,626 (100.0) | | |
| — Myocardial infarction | 995 (13.0) | 0 (0.0) | 995 (37.9) | | |
| — Stroke | 809 (10.6) | 0 (0.0) | 809 (30.8) | | |
| — All-cause death | 1,641 (21.5) | 0 (0.0) | 1,641 (62.5) | | |
| Heart failure[c] | 1,544 (20.2) | 529 (10.6) | 1,015 (38.7) | | |

*Notes.* Continuous variables are mean (SD) unless marked median [IQR]; categorical variables are n (%). SMD is the standardized mean difference between the No-MACE and MACE groups, with |SMD| > 0.1 taken to indicate non-trivial imbalance. Missingness is reported for the observed cohort; analyses used multiple imputation with 20 imputed datasets (Methods). aNot imputed and not a model input, shown for description only. bDiagnosis code, medication, or laboratory evidence of diabetes. cTracked separately and not a component of MACE.

**Supplementary Table S2. Antolini's time-dependent concordance (prespecified secondary discrimination)**

Panels A and B are computed on the **same n = 6,728 intersection cohort** as the primary analysis, so this measure and the primary measure share one denominator as well as one estimation standard. Antolini's concordance uses all **2,051 post-landmark events** rather than the 772 falling in the three-year window. Panels A and B are the prespecified secondary analysis; Panel C reports a different estimand and is retained only for reference.

**Panel A. Landmark-Antolini across the ladder, estimation-matched (primary specification)**

| Representation | Landmark-Antolini C | ΔC vs clinical-only reference | 95% CI |
|---|---|---|---|
| Clinical-only reference | 0.7092 | — | — |
| + average adherence | 0.7099 | +0.0007 | [−0.0005, +0.0018] |
| Full engineered summary | 0.7225 | +0.0133 | [+0.0084, +0.0186] |
| Fill sequence | 0.7355 | +0.0262 | [+0.0191, +0.0333] |

**Panel B. Fill sequence versus the full engineered summary, under each estimation setup**

| Comparison | C, fill sequence | C, engineered summary | ΔC | 95% CI | P(ΔC > 0) |
|---|---|---|---|---|---|
| vs engineered summary, estimation-matched (primary) | 0.7355 | 0.7225 | +0.0130 | [+0.0075, +0.0187] | >0.999 |
| vs engineered summary, time-updated (sensitivity) | 0.7355 | 0.7246 | +0.0108 | [+0.0054, +0.0163] | >0.999 |

**Panel C. Full-cohort Antolini's C, all events from the time origin (different estimand; reference only)**

| Variant | Antolini's C |
|---|---|
| V5, full engineered summary | 0.7518 |
| V4, adherence + regimen change | 0.7448 |
| V2, class indicators at t | 0.7399 |
| V3, class indicators at baseline | 0.7356 |
| V1, average adherence | 0.7294 |
| L0, clinical-only reference | 0.7286 |
| M1, fixed 180-day landmark[a] | 0.7267 |

*Notes.* Panels A and B are the prespecified secondary measure and use the same landmark estimation as the primary analysis; the time-updated row of Panel B is the corresponding sensitivity analysis. Panel C is not comparable: it integrates over the full follow-up for models estimated on the full record, a different estimand on a different risk set, and is retained only because earlier versions reported it. The fill-sequence model is absent from Panel C because it is trained on the landmark task. The increment for average adherence in Panel A does not exclude zero on this measure, consistent with the near-null increment on the primary measure. aHarrell's C, shown for reference only. P(ΔC > 0) is the share of paired bootstrap resamples exceeding zero, not a null-hypothesis P value.

##### Supplementary Table S3. Representation × algorithm factorial

Dynamic-landmark C (s = 365 d, +3 y), 5-fold cross-validation × 20 imputations. Every Cox and gradient-boosted cell is computed on the full landmark cohort (n = 6,918, 817 events). The two sequence-model cells cannot be: that model requires at least one mapped fill in the first year and therefore scores 6,822 of those patients, and the values shown are its paired-comparison estimates on the intersection cohort (n = 6,728). Absolute concordance differs by at most 0.0030 across these denominators and no ranking is affected. Every cell shares the same baseline covariates.

| Representation | n features | Linear (Cox) | Nonlinear (GBM) | Sequence model |
|---|---|---|---|---|
| Baseline (L0) | 22 | 0.7112 | 0.7180 | — |
| + average adherence, PDC (V1) | 23 | 0.7125 | 0.7204 | — |
| + regimen dynamics (V4) | 27 | 0.7257 | 0.7327 | — |
| + full engineered summary (V5) | 38 | 0.7303 | 0.7362 | — |
| Fill sequence, order discarded[a] | — | — | — | 0.7284 |
| Fill sequence, order retained (M4b) | — | — | — | 0.7396 |

*Notes.* The ordered sequence exceeded the best tabular cell (0.7396 vs 0.7362), and discarding order brought C down to the engineered summary level (0.7284, against 0.7273 for the summary on the same intersection cohort). aAn unordered mean-pooled (“bag-of-fills”) variant of the sequence model; it is a neural model, not a gradient-boosting model, and is reported in a separate row of the sequence- model column. Its Cox and gradient-boosted cells use the full landmark cohort rather than the all-model intersection cohort, so their absolute values differ from the main ladder by up to 0.0030 (V5: 0.7303 here against 0.7273 in Table S5). Because the sequence cells do not share that denominator, the sequence-versus- tabular contrast in this table should be read as an ordering rather than as an exact difference; the estimation-matched contrast on a common denominator is +0.0123 (Table S16). This table is the only presentation of the factorial.

**Supplementary Table S4. Fill-volume confound**

Paired bootstrap, B = 1000, intersection cohort n = 6,822.

| Model or comparison | C or ΔC | 95% CI |
|---|---|---|
| Volume-only model (fill and encounter counts) | C = 0.7174 | — |
| Engineered summary + raw fill count | C = 0.7288 | — |
| Nonlinear gradient boosting on the engineered features[a] | C = 0.7362 | — |
| Fill sequence − volume-only | ΔC = +0.0217 | [+0.0127, +0.0310] |
| Fill sequence − (engineered summary + raw fill count) | ΔC = +0.0103 | [+0.0021, +0.0188] |
| Fill sequence − engineered summary | ΔC = +0.0118 | [+0.0037, +0.0201] |
| Engineered summary − volume-only | ΔC = +0.0099 | [+0.0039, +0.0158] |

*Notes.* The engineered summary already contains fill rate, distinct-class count, and regimen-change counts, and the baseline already contains prior-year encounters, so the fill-sequence advantage persists after these adjustments. Every arm is estimated on the day-365 snapshot, matching the primary analysis. This denominator (n = 6,822) is larger than that of Table S5 (n = 6,728) because it requires valid predictions from a smaller set of models. aCarried over from Table S3 for context and computed there on the full landmark cohort (n = 6,918); it is not part of the paired comparison in this table.

#### Supplementary Table S5. Reclassification and clinical utility across the ladder

Concordance is the dynamic-landmark C (s = 365 d, +3 y) on the intersection cohort (n = 6,728), so that the column is directly comparable with the primary ladder. The reclassification, net-benefit, and Brier columns are pooled over 20 imputations on each variant's own landmark cohort, 6,918 for the Cox representations and 6,822 for the fill sequence, because they do not require a common denominator. All summary representations are estimated on the day-365 snapshot, matching the primary analysis. Rows are ordered by C.

| **Representation** | **C** | **Cal slope** | **IDI vs adherence** | **IDI vs summary** | **NRI@15% vs summary** | **NB@7.5%** | **NB@15%** | **NB@25%** | **IBS 5 y** |
|---|---|---|---|---|---|---|---|---|---|
| Fill sequence | **0.7396** | **0.905** | **+0.0344** | **+0.0245** | **+0.0201** | **+0.0614** | **+0.0323** | +0.0073 | 0.0807 |
| Full engineered summary | 0.7273 | 0.893 | +0.0102 | reference | reference | +0.0586 | +0.0315 | +0.0127 | **0.0789** |
| Adherence + regimen change | 0.7246 | 0.902 | +0.0066 | −0.0036 | −0.0105 | +0.0575 | +0.0305 | +0.0110 | 0.0789 |
| Class indicators at baseline | 0.7188 | 0.889 | +0.0039 | −0.0062 | −0.0106 | +0.0561 | +0.0306 | +0.0116 | 0.0791 |
| Class indicators at landmark | 0.7186 | 0.892 | +0.0050 | −0.0052 | −0.0217 | +0.0573 | +0.0293 | +0.0113 | 0.0789 |
| Average adherence | 0.7131 | 0.894 | reference | −0.0102 | −0.0359 | +0.0557 | +0.0279 | +0.0125 | 0.0790 |
| Clinical-only reference | 0.7113 | 0.892 | −0.0009 | −0.0111 | −0.0463 | +0.0554 | +0.0267 | **+0.0128** | 0.0790 |

*Notes.* The best value in each column is shown in bold; where a column ties, the highest-ranked row is bolded. For the calibration slope the best value is the one closest to 1.0. A calibration slope below 1.0 indicates over-confidence. IDI and NRI[3] are reported against both the adherence-only and the full engineered summary references, each estimated on the same landmark task; net benefit is from decision-curve analysis at the stated threshold probability.[2] Before recalibration the fill-sequence model overpredicts most in the top decile, although its calibration slope is the closest to 1.0, and it gives the least net benefit at the 25% threshold, where its wider probability spread produces more false positives; flexible recalibration removes this (Table S6). The clinical-only reference and the adherence-only model are nearly identical on IDI (a difference of −0.0009). The fixed 180-day landmark model is omitted because it conditions on a differently constructed eligible population.

**Supplementary Table S6. Recalibration diagnostics**

Leave-one-fold-out recalibration of the fill-sequence model. Calibration and net-benefit metrics are computed on the 6,822 patients the model scores; the concordance it shares with Table S5 is on the 6,728-patient intersection cohort.

| Metric | M4b (uncalibrated) | M4b · Platt | M4b · isotonic |
|---|---|---|---|
| Calibration slope | 0.905 | **0.991** | 0.905[a] |
| Decile-10 calibration gap (predicted − observed) | +0.140 | +0.087 | **+0.0005** |
| IBS at 5 y | 0.0807 | **0.0791** | —[b] |
| NB @ 7.5% | +0.0614 | +0.0611 | **+0.0644** |
| NB @ 15% | +0.0323 | +0.0325 | **+0.0338** |
| NB @ 25% | +0.0073 | +0.0098 | **+0.0146** |
| NRI @ 15% vs V5 | +0.0201 | **+0.0202** | −0.0006 |
| IDI vs V5 | **+0.0245** | +0.0059 | −0.0230 |

*Notes.* The best value in each row is shown in bold. Recalibration preserves patient ranking, so discrimination is identical to Table S5 by construction. The mean leave-one-fold-out Platt shrinkage factor was $\beta = 0.906$ (SD 0.026) across 100 folds, indicating uniform, low-variance over-confidence. Platt scaling restores the slope from 0.905 to 0.991. Isotonic recalibration gives near-perfect decile calibration and the highest net benefit at every threshold but compresses the cross-variant probability scale, lowering its IDI and NRI. The main text reports the Platt-recalibrated model. aIsotonic recalibration remaps probabilities rather than the linear-predictor scale, so the slope statistic is unchanged. bNot computed: the isotonic map is fit at a single horizon.

**Supplementary Table S7. Temporal validation (train pre-2018, test 2018 or later)**

The split assigns patients with an index year before 2018 to training (n = 5,689; 82.2% of the landmark cohort; 636 events in the horizon window) and those indexed in 2018 or later to testing (n = 1,229; 17.8%; 181 events). Supplementary Table S21 reports the same analysis at earlier, better-balanced cutoffs.

**Panel A. Concordance in the later era, by variant**

| Variant | n test | Events | Temporal C (2018+) | Random-split C | Difference |
|---|---|---|---|---|---|
| L0 | 1,229 | 181 | 0.7495 | 0.7146 | +0.0349 |
| V1 | 1,229 | 181 | 0.7501 | 0.7164 | +0.0337 |
| V5 | 1,229 | 181 | 0.7543 | 0.7314 | +0.0229 |
| M4b | 1,207 | 180 | 0.7795 | 0.7396[a] | +0.0399 |

**Panel B. Within-era paired comparison on the patients common to both models (n = 1,207; 180 events)**

| Model | C | ΔC | 95% CI | P(ΔC > 0) |
|---|---|---|---|---|
| M4b, fill sequence | 0.7825 | +0.0298 | [+0.0077, +0.0517] | 0.997 |
| V5, full engineered summary | 0.7528 | reference | — | — |

*Notes.* Every variant scores higher in the later era than under random splitting, so the 2018+ case mix was easier to discriminate and absolute era concordance is inflated; the comparison of interest is the within-era paired difference in Panel B. That difference is wider than the cross-validated +0.0123 and shows no evidence of era-related degradation. The interval is wide because the test set is small; the absence of degradation holds at every cutoff in Table S21 for the summary representations; the fill-sequence comparison exists only at the 2018 split. aFor the Cox variants the random-split reference is size-matched to the temporal test set; for M4b it is the production cross-validated estimate and is not size-matched, so the M4b difference is not comparable in magnitude with the Cox rows. Panel A reports the imputation-pooled concordance and Panel B the mean over paired bootstrap resamples of the same patients, which is why M4b appears as 0.7795 in Panel A and 0.7825 in Panel B; the two estimators differ in the same way and by a similar amount as in Table S19.

#### Supplementary Table S8. Subgroup discrimination, fill sequence versus engineered summary

All subgroups at all three horizons, landmark s = 365 d, B = 1000. The comparator is the estimation-matched landmark engineered summary, matching the primary analysis. Subgroup sizes are fixed across horizons; event counts are those falling within each horizon window.

| Horizon | Subgroup | n | Events | C, fill sequence | C, engineered summary | ΔC | 95% CI | P(ΔC > 0) |
|---|---|---|---|---|---|---|---|---|
| +1 y | Adherence, low | 2,243 | 121 | 0.7402 | 0.7199 | **+0.0203** | [+0.0016, +0.0400] | 0.984 |
| +1 y | Adherence, medium | 2,322 | 72 | 0.7620 | 0.7395 | +0.0225 | [−0.0083, +0.0528] | 0.918 |
| +1 y | Adherence, high | 2,257 | 81 | 0.7745 | 0.7847 | −0.0102 | [−0.0345, +0.0125] | 0.183 |
| +1 y | Complexity, 1 class | 5,081 | 191 | 0.7556 | 0.7414 | +0.0141 | [−0.0035, +0.0319] | 0.942 |
| +1 y | Complexity, 2 classes | 1,134 | 44 | 0.7936 | 0.7863 | +0.0074 | [−0.0228, +0.0359] | 0.693 |
| +1 y | Complexity, ≥3 classes | 607 | 39 | 0.6846 | 0.7115 | −0.0268 | [−0.0691, +0.0159] | 0.101 |
| +3 y | Adherence, low | 2,243 | 309 | 0.7343 | 0.7145 | **+0.0198** | [+0.0079, +0.0320] | >0.999 |
| +3 y | Adherence, medium | 2,322 | 263 | 0.7450 | 0.7225 | **+0.0225** | [+0.0071, +0.0373] | 0.996 |
| +3 y | Adherence, high | 2,257 | 243 | 0.7396 | 0.7453 | −0.0058 | [−0.0181, +0.0070] | 0.177 |
| +3 y | Complexity, 1 class | 5,081 | 584 | 0.7422 | 0.7269 | **+0.0153** | [+0.0053, +0.0249] | 0.998 |
| +3 y | Complexity, 2 classes | 1,134 | 137 | 0.7537 | 0.7464 | +0.0073 | [−0.0078, +0.0230] | 0.834 |
| +3 y | Complexity, ≥3 classes | 607 | 94 | 0.6823 | 0.6916 | −0.0093 | [−0.0358, +0.0166] | 0.246 |
| +5 y | Adherence, low | 2,243 | 445 | 0.7351 | 0.7177 | **+0.0174** | [+0.0069, +0.0286] | >0.999 |
| +5 y | Adherence, medium | 2,322 | 391 | 0.7364 | 0.7142 | **+0.0223** | [+0.0092, +0.0353] | >0.999 |
| +5 y | Adherence, high | 2,257 | 383 | 0.7431 | 0.7452 | −0.0021 | [−0.0125, +0.0082] | 0.347 |
| +5 y | Complexity, 1 class | 5,081 | 873 | 0.7385 | 0.7233 | **+0.0152** | [+0.0070, +0.0230] | >0.999 |
| +5 y | Complexity, 2 classes | 1,134 | 200 | 0.7554 | 0.7455 | +0.0099 | [−0.0033, +0.0228] | 0.929 |
| +5 y | Complexity, ≥3 classes | 607 | 146 | 0.6869 | 0.6998 | −0.0128 | [−0.0346, +0.0071] | 0.108 |

*Notes.* Differences whose confidence interval excludes zero are shown in bold. Adherence tertiles are defined on PDC at the landmark; complexity is the number of distinct antihypertensive classes dispensed by the landmark. The advantage concentrates in the low-adherence and single-class strata and is absent where the engineered summaries already perform well. In no subgroup does the interval lie entirely below zero. These are 18 prespecified strata examined without multiplicity adjustment and are interpreted as exploratory.

#### Supplementary Table S9. Equity and fairness audit

Discrimination and calibration by subgroup for the clinical-only reference and the fill-sequence model. Cohort sizes differ slightly between models because each requires valid predictions from its own fit.

| Variant | Subgroup | n | Events | C | Cal slope | O/E |
|---|---|---|---|---|---|---|
| L0 | White | 5,805 | 718 | 0.7079 | 0.9008 | 0.8368 |
| M4b | White | 5,725 | 716 | 0.7351 | 0.8969 | 0.7285 |
| L0 | Black | 720 | 73 | 0.7159 | 1.1367 | 0.8316 |
| M4b | Black | 713 | 73 | 0.7347 | 0.9769 | 0.7287 |
| L0 | Other or Unknown | 393 | 26 | 0.6868 | 1.0323 | 0.8161 |
| M4b | Other or Unknown | 384 | 26 | 0.7096 | 0.8946 | 0.6837 |
| L0 | Age <65 y | 4,876 | 451 | 0.7029 | 1.0478 | 0.8625 |
| M4b | Age <65 y | 4,799 | 450 | 0.7463 | 1.0174 | 0.7349 |
| L0 | Age ≥65 y | 2,042 | 366 | 0.6603 | 0.7804 | 0.8729 |
| M4b | Age ≥65 y | 2,023 | 365 | 0.6779 | 0.8016 | 0.8250 |
| L0 | Male | 3,497 | 469 | 0.7067 | 0.9156 | 0.8423 |
| M4b | Male | 3,422 | 467 | 0.7312 | 0.9552 | 0.7503 |
| L0 | Female | 3,421 | 348 | 0.7085 | 0.9437 | 0.8389 |
| M4b | Female | 3,400 | 348 | 0.7362 | 0.8599 | 0.7096 |
| L0 | Diabetes | 2,341 | 338 | 0.7143 | 0.8892 | 0.8239 |
| M4b | Diabetes | 2,322 | 338 | 0.7487 | 0.9047 | 0.7307 |
| L0 | No diabetes | 4,577 | 479 | 0.7037 | 0.9420 | 0.8470 |
| M4b | No diabetes | 4,500 | 477 | 0.7247 | 0.8946 | 0.7259 |

##### Cross-race calibration-slope gap (White − Black), all four variants

| Variant | Cal slope, White | Cal slope, Black | Gap |
|---|---|---|---|
| L0, clinical-only reference | 0.9008 | 1.1367 | −0.2358 |
| V1, average adherence | 0.9101 | 1.1726 | −0.2625 |

*continued on next page*

| Variant | Cal slope, White | Cal slope, Black | Gap |
|---|---|---|---|
| V5, full engineered summary | 0.9243 | 1.0342 | −0.1099 |
| M4b, fill sequence | 0.8969 | 0.9769 | −0.0800 |

*Notes.* A calibration slope below 1.0 indicates over-confidence; an observed-to-expected ratio of 1.0 is ideal. Representations carrying more medication information narrow the cross-race calibration gap monotonically from V1 through M4b. Discrimination is closely similar between men and women in both models, and the richer representation raises it for both. The advantage of the fill-sequence model is smallest among patients aged 65 years or older, the hardest stratum for every model. Confidence intervals are not shown because event counts as low as 26 would render them uninformative; these estimates are descriptive, and a non-significant difference would not establish equivalent performance. Race categories follow the source electronic health record.

**Supplementary Table S10. Eight-feature deployable clinical score**

A Cox model fit directly to MACE on eight forward-selected features (six non-medication variables and two medication-derived). Pooled 5-fold cross-validated C = 0.7242 (between-imputation SD 0.0003) at landmark 365 d, +3 y, 20 imputations.

| **Feature** | **Hazard ratio** | **95% CI** |
|---|---|---|
| Prior stroke | 3.255 | [2.732, 3.879] |
| Prior coronary artery disease | 2.071 | [1.824, 2.351] |
| Number of drug classes | 1.199 | [1.154, 1.245] |
| Age, per year | 1.031 | [1.027, 1.035] |
| Encounters in prior year, per visit | 1.003 | [1.002, 1.004] |
| eGFR, per mL/min/1.73 $m^2$ | 0.991 | [0.989, 0.993] |
| HDL cholesterol, per mg/dL | 0.992 | [0.989, 0.994] |
| On thiazide-type diuretic | 0.623 | [0.547, 0.709] |

*Notes.* The score (C = 0.7242) sits between the V3 and V4 rungs, below V5 (0.7273) and M4b (0.7396). It recovers approximately 46% of the fill-sequence model's gain over the clinical-only reference, (0.7242 − 0.7113)/(0.7396 − 0.7113), and is not a substitute for the full model; its value is deployability and directly interpretable hazard ratios. Six of the eight features are standard risk factors. The share of the fill-sequence signal the transparent surrogate cannot reproduce is approximately 18% (Supplementary Note 1).

**Supplementary Table S11. PDC definition sensitivity (exposure vs gap-adjusted adherence)**

Prespecified head-to-head comparison of the exposure PDC used in the main paper and a gap-adjusted adherence PDC, using the same cross-validation grid and intersection cohort.

| Comparison | ΔC | 95% CI |
|---|---|---|
| Adherence PDC − exposure PDC (V1) | −0.00006 | [−0.00070, +0.00070] |
| Both PDC definitions − exposure PDC | +0.00008 | [−0.00080, +0.00050] |

*Notes.* Values in this table alone are reported to five decimal places because the differences are smaller than the rounding unit used elsewhere. The per-patient linear-predictor correlation between the two definitions was r = 0.9997, so the small increment from average adherence is not an artifact of the PDC definition. A subgroup analysis restricted to patients with heavy gap adjustment showed no informative-of-survival artifact. The exposure PDC, which follows the Medicare Star Ratings convention, is retained as the main-paper variable.

**Supplementary Table S12. Risk classification and event capture at screening thresholds**

Common intersection cohort n = 6,822.

| Threshold | Model | Sensitivity | PPV | Lift |
|---|---|---|---|---|
| Top 5% | L0 | 18.6% | 44.4% | 3.71 |
| Top 5% | M4b | 18.0% | 42.8% | 3.58 |
| Top 10% | L0 | 29.9% | 35.6% | 2.98 |
| Top 10% | V1 | 30.3% | 36.1% | 3.02 |
| Top 10% | V5 | 30.7% | 36.6% | 3.07 |
| Top 10% | M4b | 30.8% | 36.7% | 3.08 |
| Top 25% | L0 | 52.5% | 25.1% | 2.10 |
| Top 25% | V5 | 54.6% | 26.1% | 2.18 |
| Top 25% | M4b | 55.9% | 26.7% | 2.23 |

*Notes.* Sensitivity is the share of all MACE in the cohort that occur among flagged patients; PPV is events per 100 flagged patients; lift is PPV divided by the base rate. Outreach to the highest-risk 10% of 1,000 patients (approximately 119 events) captures 35.6 events under L0, 36.1 under V1, 36.6 under V5, and 36.7 under M4b. At the extreme tail the fill-sequence model does not improve on the clinical-only reference; the fill-sequence advantage is larger in the broader top-25% band and in the low-adherence stratum (Table S8).

**Supplementary Table S13. Multicollinearity of the engineered summary (V5)**

Variance inflation factors for the engineered summary features at the day-365 landmark, computed on the 6,918 patients alive at that landmark.

| Feature | VIF |
|---|---|
| Distinct drug classes | 5.95 |
| Longest coverage gap | 5.70 |
| Exposure PDC | 5.59 |
| Cumulative add-on count | 5.07 |
| Days since last fill | 4.64 |
| Fill rate | 2.37 |
| Mean dose intensity | 2.13 |
| Current dose intensity | 2.04 |
| Cumulative switch count | 1.76 |
| Seven drug-class indicators[a] | 1.08 to 1.35 |

*Notes.* No feature reaches the conventional VIF threshold of 10, and four lie between 5 and 6. Because these four adherence and regimen-dynamics features are mutually intercorrelated, the main text interprets medication contributions by conceptual block rather than as individual hazard ratios. Discrimination depends only on the ranking of the linear predictor and is unaffected. aRange across the seven indicators, reported jointly.

**Supplementary Table S14. Deployment decomposition, snapshot baseline, and external benchmark**

Dynamic-landmark C (s = 365 d, +3 y) for the data-availability ladder and reference models. The four data-availability rows (demographics only, pharmacy-only sequence model, clinical-only reference, full fill-sequence model) are paired-bootstrap estimates (B = 1000) on the 6,822 patients the fill-sequence model can score, the denominator of main-text Figure 5. The remaining rows are context and are not part of that paired comparison: the sequence-only model is estimated on the 6,728-patient intersection cohort, the two snapshot rows are imputation-pooled, and the PREVENT row is the imputation-pooled benchmark on the 6,918 patients alive at the landmark.

| **Model** | **Inputs** | **C** |
|---|---|---|
| Demographics only | Age, sex, race | 0.6210 |
| Sequence only | Fill timeline, no covariates | 0.6510 |
| Pharmacy-only sequence model | Demographics + fill history | 0.6876 |
| Clinical-only reference (L0) | Demographics + laboratory + blood pressure | 0.7099 |
| Full fill-sequence model (M4b) | All inputs | 0.7391 |
| Last-fill snapshot Cox, s = 365 d | Most recent fill + covariates | 0.7208 |
| Last-fill snapshot Cox, s = 180 d | Most recent fill + covariates | 0.7261 |
| AHA PREVENT base equation[a] | Sex-specific 10-year ASCVD model | 0.6504 |

*Notes.* Adding fill history to demographics raised concordance by 0.0666, three quarters of the 0.0888 that laboratory data close, but did not reach the laboratory-based model: pharmacy fills complement rather than substitute for laboratory assessment. Fill history alone reached 0.6510 on the intersection cohort, above demographics alone (0.6210) though not on the same denominator. The gap between the last-fill snapshot and the full engineered summary is the headroom the sequence representation targets. aReported as external context only; its lower value partly reflects a mismatch in outcome and horizon (10-year atherosclerotic cardiovascular disease vs 3-year MACE including all-cause death), so it is not a like-for-like comparison.

#### Supplementary Table S15. Characteristics of engineered medication-history features

Engineered medication-history features, observed (not imputed).

| Feature | Overall | No MACE | MACE | Missing, n (%) | SMD |
|---|---|---|---|---|---|
| **Panel A. Patient-level engineered features at the 1-year landmark** (N = 6,918 patients) | | | | | |
| Exposure PDC, 0 to 1 | 0.67 (0.31) | 0.68 (0.31) | 0.66 (0.32) | 0 | 0.047 |
| Fill rate, fills per 30 days | 0.52 (0.45) | 0.49 (0.42) | 0.59 (0.50) | 0 | −0.209 |
| Distinct antihypertensive classes, n | 2 [1, 2] | 2 [1, 2] | 2 [1, 3] | 0 | −0.411 |
| Cumulative class add-ons, n | 1 [0, 2] | 0 [0, 2] | 1 [0, 3] | 0 | −0.329 |
| Cumulative class switches, n | 0 [0, 0] | 0 [0, 0] | 0 [0, 0] | 0 | −0.084 |
| Mean dose intensity across fills (HDD) | 0.64 (0.63) | 0.61 (0.52) | 0.71 (0.84) | 0 | −0.152 |
| Current dose intensity (HDD) | 0.64 (0.74) | 0.62 (0.64) | 0.70 (0.94) | 0 | −0.108 |
| Longest coverage gap, days | 56 [7, 180] | 50 [7, 179] | 68 [7, 186] | 0 | −0.072 |
| Days since last fill | 72 [23, 230] | 73 [23, 230] | 66 [21, 228] | 0 | 0.021 |
| *Drug class active at landmark*, n (%) | | | | | |
| ACE inhibitor | 1,475 (21.3) | 1,022 (21.5) | 453 (21.0) | 0 | 0.011 |
| Angiotensin-receptor blocker | 721 (10.4) | 507 (10.6) | 214 (9.9) | 0 | 0.024 |
| Calcium-channel blocker | 852 (12.3) | 538 (11.3) | 314 (14.6) | 0 | −0.098 |
| Beta-blocker | 1,388 (20.1) | 827 (17.4) | 561 (26.0) | 0 | −0.210 |
| Thiazide-type diuretic | 1,130 (16.3) | 846 (17.8) | 284 (13.2) | 0 | 0.127 |
| Other diuretic | 587 (8.5) | 308 (6.5) | 279 (12.9) | 0 | −0.219 |
| Other antihypertensive | 87 (1.3) | 49 (1.0) | 38 (1.8) | 0 | −0.063 |
| **Panel B. Fill-level token features** (223,201 fills across 7,550 patients) | | | | | |
| Days supplied per fill | 90 [30, 90] | 90 [30, 90] | 34 [30, 90] | 0 | 0.080 |
| Dose intensity per fill (HDD) | 0.64 (1.01) | 0.61 (0.74) | 0.69 (1.29) | 754 (0.3) | −0.067 |
| Early refill, n (%) | 80,853 (36.2) | 47,794 (36.9) | 33,059 (35.3) | 0 | 0.033 |
| Calendar year of fill | 2016 [2014, 2018] | 2016 [2014, 2018] | 2016 [2014, 2018] | 0 | 0.206 |
| *Drug class of fill*, n (%) | | | | | |
| ACE inhibitor | 50,176 (22.5) | 33,028 (25.5) | 17,148 (18.3) | 0 | 0.174 |
| Angiotensin-receptor blocker | 27,848 (12.5) | 17,995 (13.9) | 9,853 (10.5) | 0 | 0.103 |
| Calcium-channel blocker | 32,922 (14.7) | 19,138 (14.8) | 13,784 (14.7) | 0 | 0.001 |
| Beta-blocker | 56,904 (25.5) | 29,739 (23.0) | 27,165 (29.0) | 0 | −0.138 |
| Thiazide-type diuretic | 36,610 (16.4) | 25,909 (20.0) | 10,701 (11.4) | 0 | 0.236 |
| Other diuretic | 30,277 (13.6) | 13,930 (10.8) | 16,347 (17.5) | 0 | −0.193 |
| Other antihypertensive | 5,408 (2.4) | 2,232 (1.7) | 3,176 (3.4) | 0 | −0.106 |

*Notes.* Continuous features are mean (SD) or median [IQR] as marked; binary features are n (%). SMD is the standardized mean difference between the No-MACE and MACE groups, with |SMD| > 0.1 taken to indicate non-trivial imbalance. MACE is the 3-point composite over follow-up, as in Table 1.

*Panel A.* Full one-year landmark cohort (N = 6,918; No MACE n = 4,763; post-landmark MACE n = 2,155), evaluated at the last 30-day interval boundary at or before the landmark, which is day 360, among patients event-free at day 365. HDD is normalized; 0 denotes no dosed fill.

*Panel B.* One row per fill, restricted to the 223,201 fills read by the sequence model, that is, fills mapped to the seven drug classes (median 17 fills per patient [IQR 6, 39]). These fills belong to 7,550 of the 7,625 patients in the analytic cohort; the remaining 75 have no fill mapping to a modeled class anywhere in the record and so contribute no tokens. The full record contains 306,501 fills including pre-index and unmapped fills. Fills are nested within patients and the split is by the patient's MACE status, so the standardized differences in this panel are not independent observations. Combination products contribute to more than one drug class, so the class percentages sum to more than 100%. The median days supplied differs between the groups (34 against 90) while the quartiles do not, because fills in the MACE group draw more heavily on 30-day supplies, consistent with their higher share of beta-blocker and other-diuretic fills; the standardized difference is computed on means and is small.

**Supplementary Table S16. Time-updated estimation, sensitivity analysis**

Paired bootstrap, B = 1000, intersection cohort n = 6,728 with 772 events in the horizon window. The primary analysis estimates every summary representation as an ordinary Cox model on the day-365 feature snapshot against the post-landmark outcome, matching the target on which the fill-sequence model is trained. This table reports the sensitivity analysis in which the same rungs are instead estimated as counting-process models on the full longitudinal record and then evaluated with the linear predictor frozen at day 365.

**Panel A. Effect of the estimation setup, by time-varying medication content**

| Representation | Medication features | Landmark C (primary) | Time-updated C (sensitivity) | ΔC (time-updated − landmark) | 95% CI |
|---|---|---|---|---|---|
| L0, clinical-only reference | 0 | 0.7113 | 0.7115 | +0.0001 | [−0.0025, +0.0030] |
| V1, average adherence | 1 | 0.7131 | 0.7149 | +0.0018 | [−0.0006, +0.0046] |
| V4, adherence + regimen change | 5 | 0.7246 | 0.7269 | +0.0023 | [−0.0005, +0.0053] |
| V5, full engineered summary | 16 | 0.7273 | 0.7316 | +0.0044 | [+0.0002, +0.0089] |

**Panel B. Model comparison under each specification**

| Comparison | ΔC | 95% CI | P(ΔC > 0) |
|---|---|---|---|
| Fill-sequence − full engineered summary, estimation-matched (primary) | +0.0123 | [+0.0040, +0.0202] | >0.999 |
| Fill-sequence − full engineered summary, time-updated (sensitivity) | +0.0080 | [−0.0003, +0.0157] | 0.964 |
| Fill-sequence − adherence + regimen change, estimation-matched | +0.0150 | [+0.0062, +0.0234] | 0.999 |
| Full engineered summary − clinical-only reference, estimation-matched | +0.0159 | [+0.0090, +0.0239] | >0.999 |
| Average adherence − clinical-only reference, estimation-matched | +0.0017 | [+0.0002, +0.0032] | 0.980 |
| Average adherence − clinical-only reference, time-updated | +0.0034 | [+0.0013, +0.0055] | 0.998 |

*Notes.* "Landmark" denotes the primary estimation, a Cox model refit on the day-365 snapshot; "time-updated" denotes the counting-process estimation used in the sensitivity analysis. The time-updated specification favors the summary models, and the benefit increases monotonically with the amount of time-varying medication information a representation carries. For the clinical-only reference, which contains no medication features, the difference is null; this negative control indicates the effect arises from the medication features rather than from the estimation procedure in general. The fill-sequence model still leads under the less favorable specification. P(ΔC > 0) is the share of paired bootstrap resamples exceeding zero, not a null-hypothesis P value.

##### Supplementary Table S17. Medication-change representation

Paired bootstrap, B = 1000, intersection cohort n = 6,728 with 772 events in the horizon window. Discrimination at the primary landmark (day 365) and horizon (+3 years) under alternative representations of class switching and add-on therapy. Because that block is defined over the whole record, this re-specification was carried out under the time-updated estimation, so every value in this table, including the fill-sequence model, is estimated that way and should be read against the estimation-matched primary difference of +0.0123 only as an ordering.

###### Panel A. Absolute discrimination

| **Representation** | **Switch/add-on features** | **C** |
|---|---|---|
| Adherence + regimen change (published, V4) | Published binary counts | 0.7269 |
| Adherence + regimen change | Corrected classification | 0.7273 |
| Adherence + regimen change | Removed | 0.7276 |
| Adherence + regimen change | Continuous concurrency | 0.7291 |
| Full engineered summary (published, V5) | Published binary counts | 0.7316 |
| Full engineered summary | Corrected classification | 0.7321 |
| Full engineered summary | Continuous concurrency | 0.7333 |
| Fill sequence (M4b) | — | 0.7384 |

###### Panel B. Contrasts

| **Comparison** | **ΔC** | **95% CI** | **P(ΔC > 0)** |
|---|---|---|---|
| Removed − published (adherence summary) | +0.0007 | [−0.0003, +0.0018] | 0.907 |
| Corrected classification − published (adherence summary) | +0.0004 | [−0.0016, +0.0023] | 0.636 |
| Continuous concurrency − published (adherence summary) | +0.0022 | [−0.0007, +0.0051] | 0.933 |
| Continuous concurrency − corrected classification | +0.0018 | [−0.0001, +0.0037] | 0.973 |
| Continuous concurrency − published (full summary) | +0.0016 | [−0.0009, +0.0044] | 0.887 |
| Fill sequence − published (full summary) | +0.0068 | [−0.0021, +0.0150] | 0.928 |
| Fill sequence − continuous concurrency (full summary) | +0.0051 | [−0.0037, +0.0134] | 0.828 |

*Notes.* The confidence interval includes zero for every contrast in this table. Removing the switch and add-on features does not reduce discrimination, indicating that these counts carried no discrimination independent of the distinct-class count and refill rate already present in the summary. The continuous concurrency representation is the strongest of the three, but its interval touches zero and the ordering did not hold on the prespecified time-dependent concordance (Table S2). On this table's own estimation and denominator the fill-sequence model reaches 0.7384, so its advantage is +0.0068 (95% CI −0.0021 to +0.0150) over the published summary and +0.0051 (−0.0037 to +0.0134) over the strengthened continuous-concurrency summary; both intervals include zero. Strengthening the switch and add-on block therefore narrows the gap without closing it. The main-text ladder, which is estimation-matched, is unchanged.

##### Supplementary Table S18. Sources of within-band discrimination

Bootstrap, B = 1000, common cohort n = 6,822 with 815 events in the horizon window (all five scores are available for every patient, so the rows within a band are directly comparable). Discrimination at the primary landmark (day 365) and horizon (+3 years), computed separately within each adherence band. All scores are cross-validated out-of-fold predictions.

| **Adherence band** | **n** | **Events** | **Score** | **C (95% CI)** |
|---|---|---|---|---|
| Adherent (PDC ≥0.80) | 3,240 | 346 | Average adherence (PDC), single variable | 0.4628 (0.4322 to 0.4934) |
| Adherent (PDC ≥0.80) | 3,240 | 346 | Clinical covariates only | 0.7227 (0.6977 to 0.7488) |
| Adherent (PDC ≥0.80) | 3,240 | 346 | Clinical covariates + PDC | 0.7222 (0.6976 to 0.7483) |
| Adherent (PDC ≥0.80) | 3,240 | 346 | Fill sequence only, no covariates | 0.6499 (0.6211 to 0.6762) |
| Adherent (PDC ≥0.80) | 3,240 | 346 | Clinical covariates + fill sequence | 0.7455 (0.7220 to 0.7694) |
| Intermediate (0.50–0.80) | 1,555 | 182 | Average adherence (PDC), single variable | 0.5229 (0.4820 to 0.5638) |
| Intermediate (0.50–0.80) | 1,555 | 182 | Clinical covariates only | 0.6825 (0.6437 to 0.7201) |
| Intermediate (0.50–0.80) | 1,555 | 182 | Clinical covariates + PDC | 0.6826 (0.6431 to 0.7204) |
| Intermediate (0.50–0.80) | 1,555 | 182 | Fill sequence only, no covariates | 0.6683 (0.6294 to 0.7025) |
| Intermediate (0.50–0.80) | 1,555 | 182 | Clinical covariates + fill sequence | 0.7254 (0.6872 to 0.7589) |
| Low (PDC <0.50) | 2,027 | 287 | Average adherence (PDC), single variable | 0.5020 (0.4693 to 0.5357) |
| Low (PDC <0.50) | 2,027 | 287 | Clinical covariates only | 0.7164 (0.6851 to 0.7472) |
| Low (PDC <0.50) | 2,027 | 287 | Clinical covariates + PDC | 0.7164 (0.6845 to 0.7471) |
| Low (PDC <0.50) | 2,027 | 287 | Fill sequence only, no covariates | 0.6445 (0.6132 to 0.6745) |
| Low (PDC <0.50) | 2,027 | 287 | Clinical covariates + fill sequence | 0.7404 (0.7115 to 0.7677) |

*Notes.* This table separates the sources of the main-text within-band difference; every score shares one estimation standard. Adding average adherence to the clinical covariates changes discrimination by less than 0.0006 in every band, so the within-band null is not an artifact of the single-variable comparison. The same null holds in the low-adherence band, where average adherence spans the whole range below the threshold (mean 0.26): average adherence discriminates between bands (whole-cohort C 0.5310) but not within them. The fill sequence reaches C 0.6499 in the adherent band with no covariates, and the full model exceeds the clinical covariates by 0.0228 there, though most absolute within-band discrimination still comes from the clinical covariates.

##### Supplementary Table S19. PREVENT-anchored representation ladder

Paired bootstrap, B = 1000, common cohort n = 6,822 with 815 events in the horizon window — the same cohort as Supplementary Table S18, comprising the patients whom the fill-sequence model could score at the day-365 landmark. Folds (seed 42), imputations, penalization, and the headline metric (dynamic-landmark concordance at day 365 with a +3-year horizon) match the primary ladder. Each rung adds one term on top of the recalibrated PREVENT linear predictor, so its increment is the incremental value over the guideline calculator itself. Expands main-text Figure 2B.

###### Panel A. Discrimination by rung

| Rung | Terms added to PREVENT | Paired-bootstrap C | Imputation-pooled C | SD across 20 imputations | Range across imputations |
|---|---|---|---|---|---|
| PREVENT,[4] recalibrated | — | 0.6512 | 0.6488 | 0.0018 | 0.6459 to 0.6523 |
| + average adherence (PDC) | 1 | 0.6537 | 0.6514 | 0.0018 | 0.6483 to 0.6548 |
| + full engineered summary | 16 | 0.6829 | 0.6813 | 0.0013 | 0.6795 to 0.6834 |
| + fill-sequence score | 1 (model score) | 0.6969 | 0.6957 | 0.0011 | 0.6941 to 0.6977 |
| Fully internally fitted model (M4b), for scale | not a rung | 0.7391 | — | — | — |

###### Panel B. Incremental contrasts

| Contrast | ΔC | 95% CI | P(ΔC > 0) |
|---|---|---|---|
| + average adherence over PREVENT | +0.0025 | [−0.0003, +0.0054] | 0.953 |
| + full engineered summary over PREVENT | +0.0317 | [+0.0222, +0.0414] | >0.999 |
| + fill-sequence score over PREVENT | +0.0458 | [+0.0319, +0.0589] | >0.999 |
| Full engineered summary over average adherence | +0.0292 | [+0.0190, +0.0385] | >0.999 |
| Fill sequence over average adherence | +0.0432 | [+0.0285, +0.0570] | >0.999 |
| Fill sequence over full engineered summary | +0.0140 | [+0.0029, +0.0250] | 0.990 |
| Fully internally fitted model over PREVENT (context, not a rung) | +0.0879 | [+0.0714, +0.1057] | >0.999 |
| Fully internally fitted model over the fill-sequence rung (context) | +0.0422 | [+0.0315, +0.0532] | >0.999 |

*Notes.* This analysis tests the premise on which the antihypertensive indicator rests in guideline risk equations: in a cohort in which every patient is treated the indicator is constant and carries no information, so the question is whether the medication-taking record supplies what it cannot. Substituting average adherence does not (+0.0025, interval including zero; 4.7% of resamples favored PREVENT alone); representations preserving refill timing and regimen evolution do (+0.0317 and +0.0458, both excluding zero). The increments are larger here because PREVENT is a weaker starting point in this cohort: its coefficients are external and fixed, it was developed for a 10-year atherosclerotic endpoint, and it carries no term for prior cardiovascular disease, comorbidity, or healthcare contact. The comparison concerns ranking, not absolute risk.

Two contrasts deserve separate note. First, the fill sequence exceeds the full engineered summary by +0.0140 [+0.0029, +0.0250] against this anchor, whereas the corresponding primary internal contrast is +0.0123 [+0.0040, +0.0202]. The two are not in conflict: an external anchor that explains less of the outcome leaves more room for any medication representation, so the anchored contrast is not evidence of a larger sequence advantage, and the internally referenced primary comparison remains the basis for the paper's claim. Second, the fully internally fitted model exceeds PREVENT by +0.0879. That gap combines representation with the additional clinical content and the internal fitting of the reference model, so it is external context rather than a representation effect, and it is reported only to place the anchored rungs on a scale.

The paired-bootstrap and imputation-pooled concordances in Panel A differ by approximately 0.0020 because they are different estimators — bootstrap resamples of patients versus Rubin pooling across imputations. Main-text Figure 2B displays the paired-bootstrap values, matching the estimator used for the ladder in Figure 2A, so that the increments and the displayed endpoints come from one estimator. Each is rounded independently, so an increment may differ from the difference of two rounded endpoints by 0.0001.

*Denominators.* The three cohorts reported in this article are strictly nested. Of the 6,918 patients alive at the landmark (Supplementary Table S3), 96 could not be scored by the fill-sequence model, which requires at least one fill mapping to a modeled drug class in the first year; the remaining 6,822 form the cohort of this table, Table S18, and main-text Figure 2B. A further 94 are absent from M1, the fixed 180-day landmark Cox model, which is defined on a differently constructed eligible population, leaving the 6,728-patient all-model intersection used for the primary ladder (Figure 2A). The PREVENT equation was computable for every patient and therefore did not restrict any cohort. The PREVENT concordance of 0.65 reported in Supplementary Table S14 is a different quantity: it is the imputation-pooled value on the 6,918 patients alive at the landmark, not the paired-bootstrap value on the 6,822 scored here.

##### Supplementary Table S20. Complete-case sensitivity analysis

Five baseline measurements are incompletely recorded (systolic blood pressure 41%, body-mass index 30%, total and HDL cholesterol and creatinine-derived kidney function 13% to 17%), and the primary analysis multiply imputes them. Of the 6,918 patients alive at the landmark, **3,462 have every one of the 22 model covariates observed**; these carry 447 events in the three-year window, roughly half the 772 available in the primary analysis, so intervals here are correspondingly wider.

**Panel A. Ladder refit on complete cases with no imputation**

Each rung refit as a landmark Cox model on observed values only, single dataset, with the fold assignment, penalizer, landmark, horizon, and metric unchanged. Paired bootstrap, B = 1000, n = 3,462.

| **Representation** | **C** | **ΔC vs clinical-only reference** | **95% CI** |
|---|---|---|---|
| Clinical-only reference | 0.7482 | — | — |
| + average adherence | 0.7499 | +0.0017 | [−0.0006, +0.0042] |
| + adherence and regimen change | 0.7585 | +0.0104 | [+0.0045, +0.0163] |
| Full engineered summary | 0.7588 | +0.0106 | [+0.0021, +0.0192] |

**Panel B. All models evaluated on the complete-case subset**

Panel A refits each Cox rung on observed values alone, which the fill-sequence model and the fixed 180-day comparator cannot be. Panel B therefore takes the primary, imputation-based predictions and restricts the evaluation to complete-case patients who also carry a valid prediction from every model in the comparison. That second requirement removes a further 114 patients and 28 events, which is why Panel B is estimated on 3,348 patients with 419 events rather than the 3,462 with 447 of Panel A.

Every model trained exactly as in the primary analysis, with the paired bootstrap restricted to complete-case patients, so all models are compared on identical fully-observed patients. n = 3,348 with 419 events.

| **Comparison** | **ΔC** | **95% CI** | **P(ΔC > 0)** |
|---|---|---|---|
| Fill sequence − full engineered summary | +0.0051 | [−0.0049, +0.0150] | 0.828 |
| Fill sequence − clinical-only reference | +0.0169 | [+0.0065, +0.0280] | 0.999 |
| Full engineered summary − clinical-only reference | +0.0118 | [+0.0039, +0.0201] | 0.998 |
| Average adherence − clinical-only reference | +0.0015 | [−0.0004, +0.0033] | 0.941 |

*Notes.* Panel A tests whether the ladder is an artifact of imputation: the increment for average adherence is +0.0017 with no imputation anywhere, identical to the primary analysis. Panel B evaluates all models on fully-observed patients; the sequence advantage is preserved in direction but attenuated with an interval including zero, expected with roughly half the events, and is reported as a limitation rather than confirmation. The fill-sequence model's medication inputs are never imputed, since fills are observed dispensing events; only its baseline-covariate head shares the imputed covariates, exactly as the Cox models do. A neural model retrained from scratch on complete cases alone was not fitted. Absolute concordance is higher throughout because complete-case patients are a differently constituted population, so values should be compared within this table and not against the primary ladder.

**Supplementary Table S21. Temporal-split cutoff**

The temporal validation trains on patients with an index year before the cutoff and tests on those at or after it. The reported analysis uses 2018. This table gives the split proportions explicitly and repeats the analysis at earlier, better-balanced cutoffs. Every arm is a landmark Cox model, matching the primary estimand; 20 imputations; the random-split reference repeats each fit with a stratified random split of the same test size, so the difference isolates calendar-era drift from the effect of a smaller training set.

**Panel A. Split balance**

| Cutoff | Train n | Train % | Test n | Test % | Events, train | Events, test |
|---|---|---|---|---|---|---|
| 2015 | 3,554 | 51.4% | 3,364 | 48.6% | 400 | 417 |
| 2016 | 4,164 | 60.2% | 2,754 | 39.8% | 460 | 357 |
| 2017 | 4,853 | 70.2% | 2,065 | 29.8% | 545 | 272 |
| **2018 (reported)** | **5,689** | **82.2%** | **1,229** | **17.8%** | **636** | **181** |

**Panel B. Discrimination by cutoff**

| Cutoff | Representation | Temporal C | Random-split C | Difference (temporal − random) |
|---|---|---|---|---|
| 2015 | Clinical-only reference | 0.7314 | 0.7274 | +0.0040 |
| 2015 | + average adherence | 0.7343 | 0.7297 | +0.0046 |
| 2015 | Full engineered summary | 0.7394 | 0.7400 | −0.0006 |
| 2016 | Clinical-only reference | 0.7580 | 0.7285 | +0.0295 |
| 2016 | + average adherence | 0.7612 | 0.7298 | +0.0314 |
| 2016 | Full engineered summary | 0.7599 | 0.7382 | +0.0217 |
| 2017 | Clinical-only reference | 0.7511 | 0.7291 | +0.0220 |
| 2017 | + average adherence | 0.7545 | 0.7299 | +0.0246 |
| 2017 | Full engineered summary | 0.7556 | 0.7412 | +0.0144 |
| **2018 (reported)** | Clinical-only reference | 0.7495 | 0.7146 | +0.0349 |
| **2018 (reported)** | + average adherence | 0.7501 | 0.7164 | +0.0337 |
| **2018 (reported)** | Full engineered summary | 0.7543 | 0.7314 | +0.0229 |

*Notes.* Reported as temporal minus random-split concordance, the same sign convention as Table S7; the 2018 rows reproduce that table exactly. A positive difference indicates no evidence of degradation over calendar time. The only negative value anywhere is −0.0006 for the full engineered summary at the 2015 cutoff, indistinguishable from no change. The ordering of the representations is preserved at every cutoff in the random-split column, and at the 2015, 2017, and 2018 cutoffs in the temporal column. At the 2016 cutoff the full engineered summary (0.7599) falls 0.0013 below average adherence (0.7612) in the temporal column, a difference an order of magnitude smaller than the era effect the table is reporting, and smaller than the width of any interval reported for a contrast of this kind elsewhere in this Supplement. The reported 2018 cutoff is the least balanced of those examined and leaves the fewest test-era events, which is why the temporal interval in the main text is wide. The comparison against the fill-sequence model is available only at the 2018 cutoff, because that model was trained once under that split.

#### Supplementary Table S22. Group-based trajectory modeling of first-year coverage

**Panel A. Trajectory groups** (N = 7,625; the trajectory model is fitted on the full analytic cohort, whereas the landmark evaluation in Panel C uses the 6,728-patient intersection cohort. Incidence is measured over three years from the index date.)

| Group | Pattern | n | % | Mean posterior | Month 1 | Month 6 | Month 12 | Year-1 PDC, mean (SD) | 3-year incidence (95% CI) |
|---|---|---|---|---|---|---|---|---|---|
| 6 | Sustained high coverage | 2,723 | 35.7 | 0.927 | 0.99 | 0.96 | 0.93 | 0.97 (0.03) | 11.2 (10.0, 12.6) |
| 5 | Decline and partial recovery | 1,601 | 21.0 | 0.902 | 0.93 | 0.70 | 0.75 | 0.80 (0.11) | 10.0 (8.6, 11.7) |
| 2 | Early discontinuation | 1,430 | 18.8 | 0.979 | 0.84 | 0.00 | 0.02 | 0.18 (0.09) | 14.4 (12.5, 16.6) |
| 3 | Interruption and return | 661 | 8.7 | 0.931 | 0.90 | 0.14 | 0.53 | 0.40 (0.13) | 12.7 (10.3, 15.6) |
| 1 | Mid-year discontinuation | 638 | 8.4 | 0.974 | 0.96 | 0.57 | 0.00 | 0.45 (0.07) | 15.2 (12.4, 18.6) |
| 4 | Late discontinuation | 572 | 7.5 | 0.963 | 0.97 | 0.88 | 0.00 | 0.66 (0.11) | 11.3 (8.8, 14.4) |

**Panel B. Bayesian information criterion grid**

| Groups | Order 1 | Order 2 | Order 3 |
|---|---|---|---|
| 1 | 181,593 | 180,210 | 179,732 |
| 2 | 140,918 | 138,656 | 138,002 |
| 3 | 132,212 | 130,351 | 129,647 |
| 4 | 127,780 | 124,089 | 123,049 |
| 5 | 124,968 | 122,283 | 121,270 |
| 6 | 122,914 | 119,805 | 117,972 |

**Panel C. Discrimination**

| Model | Landmark C | Antolini C |
|---|---|---|
| Clinical-only reference | 0.7113 | 0.7092 |
| Average adherence | 0.7131 | 0.7099 |
| Trajectory group | 0.7118 | 0.7092 |
| Adherence with regimen change | 0.7246 | 0.7199 |

*continued on next page*

| Model | Landmark C | Antolini C |
|---|---|---|
| Full engineered summary | 0.7273 | 0.7225 |
| Fill sequence | 0.7396 | 0.7355 |

| Contrast | Landmark C | Antolini C |
|---|---|---|
| Trajectory group – average adherence | −0.0013 (−0.0030, 0.0004) | −0.0007 (−0.0019, 0.0005) |
| Trajectory group – clinical-only reference | 0.0004 (−0.0018, 0.0027) | −0.0000 (−0.0017, 0.0016) |
| Full engineered summary – trajectory group | 0.0155 (0.0085, 0.0231) | 0.0133 (0.0086, 0.0185) |
| Fill sequence – trajectory group | 0.0278 (0.0178, 0.0381) | 0.0263 (0.0192, 0.0337) |

*Notes.* n = 6,728 with 772 events in the three-year window and 2,051 post-landmark events for the Antolini metric. Trajectory values are expected coverage on the observed scale, not the fitted polynomial; the latent mean of a censored-normal model runs outside [0,1] and is not a coverage fraction. Selection was by Bayesian information criterion within a prespecified range of one to six groups and polynomial orders one to three, subject to each group containing at least 5% of the cohort with mean posterior assignment probability of at least 0.70; the criterion was still decreasing at the largest specification examined, so the selected solution is the best admissible one within that range rather than an interior optimum. Fold-wise assignment agreed with the full-cohort solution for 97.3% of patients. Group membership alone, without clinical covariates, gave a concordance of 0.5435 against 0.5310 for average adherence, so the classes carry marginally more prognostic signal than the average but none of it in addition to the clinical covariates. Of the variance in year-1 average adherence, 7.6% remained within groups. Three-year incidences are unadjusted; risk separates discontinuation from maintenance but does not track the amount of coverage, and part of the non-monotonicity across the three higher-coverage groups plausibly reflects treatment intensity tracking disease severity.

**Supplementary Table S23. Cause-specific outcome, held-out predictions re-scored**

| | **All-cause composite** | **Cardiovascular** | **MI or stroke** | **Uncoded causes as cardiovascular** |
|---|---|---|---|---|
| n / events | 6,728 / 772 | 6,687 / 640 | 6,677 / 613 | 6,694 / 661 |
| Clinical-only reference | 0.7113 | 0.7126 | 0.7105 | 0.7129 |
| Average adherence | 0.7131 | 0.7140 | 0.7119 | 0.7146 |
| Adherence with regimen change | 0.7246 | 0.7275 | 0.7258 | 0.7279 |
| Full engineered summary | 0.7273 | 0.7287 | 0.7267 | 0.7300 |
| Fill sequence | 0.7396 | 0.7421 | 0.7401 | 0.7428 |
| Fill sequence − full engineered summary | 0.0123 (0.0040, 0.0202) | 0.0133 (0.0050, 0.0216) | 0.0134 (0.0052, 0.0223) | 0.0128 (0.0047, 0.0211) |
| Fill sequence − clinical-only reference | 0.0283 (0.0189, 0.0382) | 0.0295 (0.0193, 0.0402) | 0.0295 (0.0194, 0.0405) | 0.0300 (0.0207, 0.0402) |
| Average adherence − clinical-only reference | 0.0017 (0.0002, 0.0032) | 0.0014 (−0.0002, 0.0031) | 0.0013 (−0.0006, 0.0032) | 0.0017 (0.0001, 0.0034) |

*Notes.* Underlying cause of death was coded for 1,446 of 1,641 deaths (88.1%), of which 506 were circulatory. Patients without a qualifying event are censored at end of follow-up; the at-risk set falls from 6,728 to 6,687 because 41 patients contribute an event to the all-cause composite that the cause-specific definition cannot evaluate: their death is ascertained after the landmark from the state death index, whose coverage does not depend on health-system contact, but their health-system follow-up had already ended within the first year, so no underlying cause is available to classify them by. The smaller falls to 6,677 and 6,694 in the other two columns arise the same way, and differ only because each definition classifies a slightly different set of deaths. The ordering of the representations is identical under all four definitions. Within the adherent band, average adherence gave a concordance of 0.4476 against cardiovascular events compared with 0.4628 against the all-cause composite, while the fill-sequence model gave 0.7540 against 0.7455; observed three-year cardiovascular incidence across thirds of fill-sequence predicted risk within the band was 2.8%, 5.7% and 19.4%. The increment for average adherence over the clinical-only reference ranges from 0.0013 to 0.0017 across the four definitions, with a lower confidence bound that crosses zero in two of them, so this increment should be read as a magnitude and not as a significance test. Models were trained on the all-cause composite, so this is an evaluation-side analysis; non-cardiovascular death is an informative censoring event, which biases absolute risk but affects between-model discrimination far less because every model faces the same censoring pattern on the same patients.

**Supplementary Table S24. Cause-specific outcome, every representation refitted**

| Representation | Refitted | Re-scored | Difference |
|---|---|---|---|
| Clinical-only reference | 0.7287 | 0.7126 | 0.0161 (0.0108, 0.0214) |
| Average adherence | 0.7299 | 0.7140 | 0.0159 (0.0107, 0.0213) |
| Trajectory group | 0.7296 | 0.7131 | 0.0165 (0.0114, 0.0218) |
| Adherence with regimen change | 0.7433 | 0.7275 | 0.0158 (0.0105, 0.0211) |
| Full engineered summary | 0.7446 | 0.7287 | 0.0159 (0.0107, 0.0213) |
| Fill sequence | 0.7566 | 0.7421 | 0.0145 (0.0075, 0.0215) |

| Contrast among refitted models | Landmark C | Antolini C |
|---|---|---|
| Fill sequence − full engineered summary | 0.0119 (0.0026, 0.0212) | 0.0154 (0.0084, 0.0227) |
| Fill sequence − clinical-only reference | 0.0279 (0.0164, 0.0398) | 0.0306 (0.0220, 0.0399) |
| Fill sequence − trajectory group | 0.0270 (0.0155, 0.0390) | 0.0299 (0.0213, 0.0391) |
| Average adherence − clinical-only reference | 0.0013 (−0.0013, 0.0039) | 0.0008 (−0.0012, 0.0029) |
| Trajectory group − average adherence | −0.0004 (−0.0025, 0.0019) | −0.0000 (−0.0017, 0.0016) |

*Notes.* n = 6,687 with 640 cause-specific events in the three-year window and 1,382 post-landmark events for the Antolini metric. Refitting used a cause-specific hazard formulation in which only the event indicator and the censoring time changed; subdistribution hazard models were not used. To contain computation, refitting used the first five imputed datasets and one training run per fold rather than the twenty imputations and three runs of the primary analysis, a reduction applied identically to every representation that reduces precision rather than introducing bias. Refitting improved every representation by a similar margin, with all intervals excluding zero and overlapping one another, so matching the training objective to the outcome is worth about 0.016 to every representation alike and raises the ladder without reordering it. This is not an argument for changing the primary outcome: the gain is uniform, and the all-cause composite remains justified by complete ascertainment, since 11.9% of deaths carry no coded underlying cause. The gain to the fill-sequence model is the smallest of the six, the direction opposite to a model riding on mortality, although the intervals overlap heavily and this is not claimed as a real difference.

### Supplementary Figures

#### Supplementary Figure S1. Calibration by decile, all variants

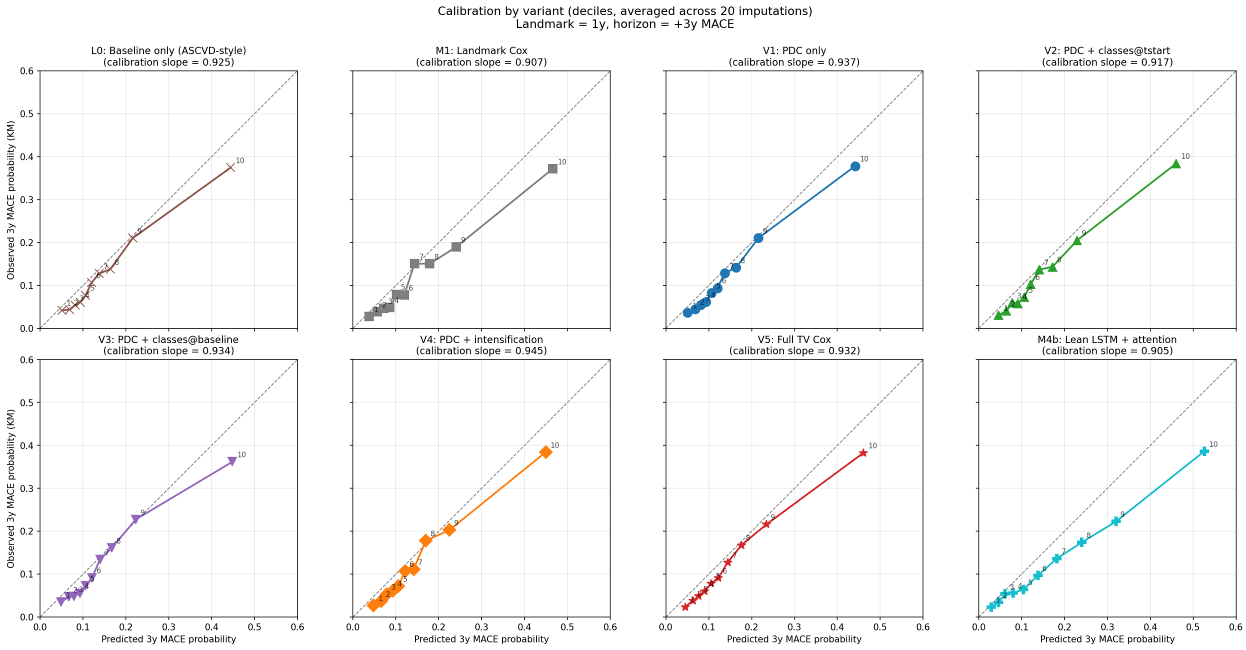

*Notes.* Predicted versus observed three-year MACE risk by decile of predicted risk for each variant. Expands main-text Figure 4A.

**Supplementary Figure S2. Decision-curve analysis across the full threshold range**

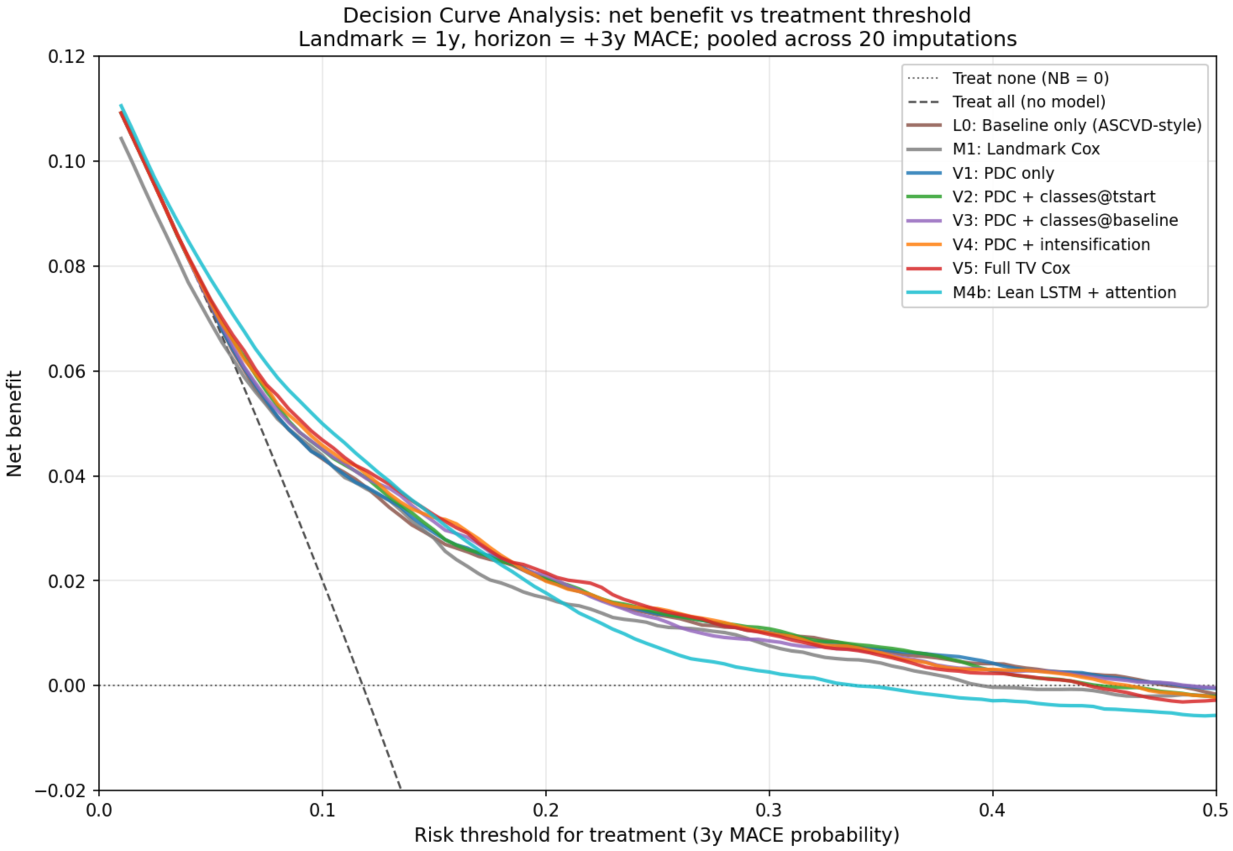

*Notes.* Net benefit versus threshold probability (0.02 to 0.40) for each variant, with treat-all and treat-none references. Expands main-text Figure 4B.

**Supplementary Figure S3. Learning curve of the fill-sequence model**

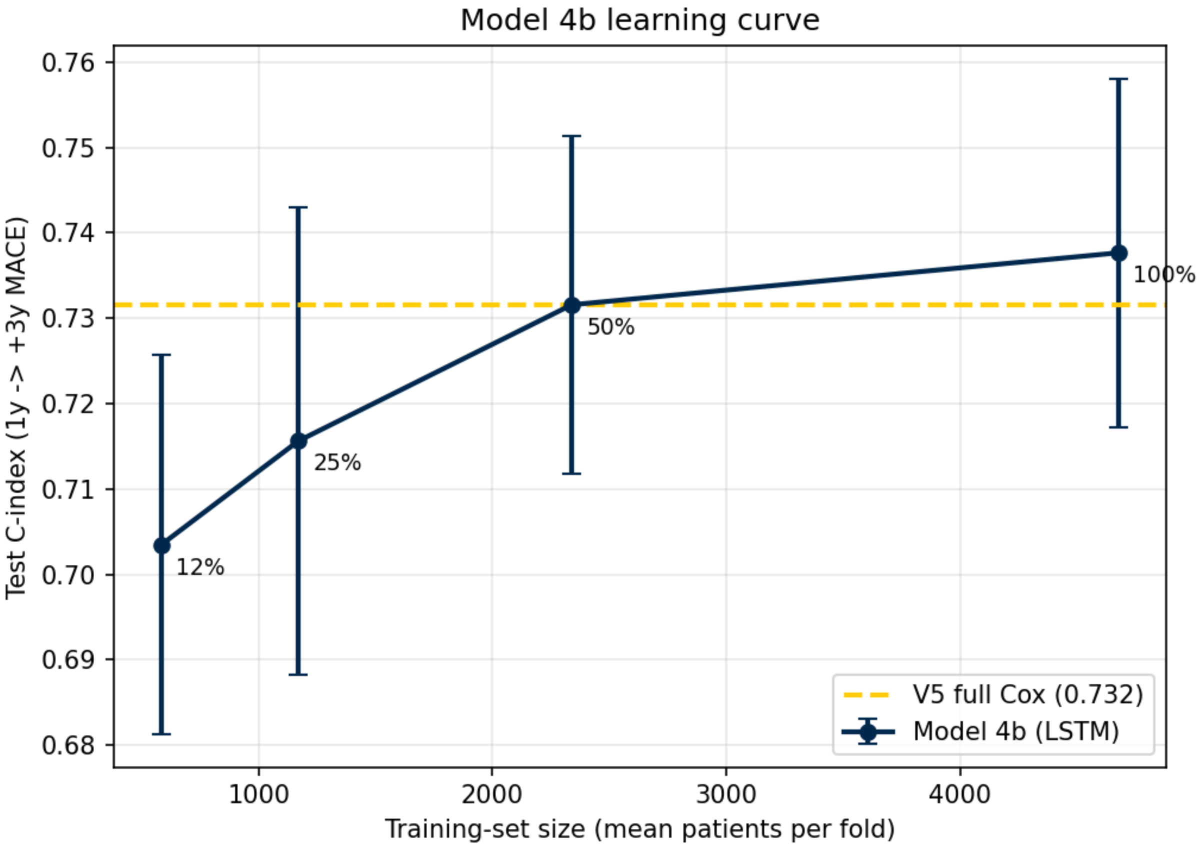

*Notes.* Held-out concordance versus training-set fraction: 0.7035 at 12.5%, 0.7156 at 25%, 0.7316 at 50%, and 0.7377 at 100%. The curve is still rising at the full sample size (the 50% to 100% increment is +0.0061, above a 0.0030 plateau threshold), so performance had not clearly plateaued at the available sample size. The 100% endpoint differs slightly from the headline production estimate because the latter also averages three random seeds across the 20 imputations.

**Supplementary Figure S4. Cohort derivation and landmark denominators**

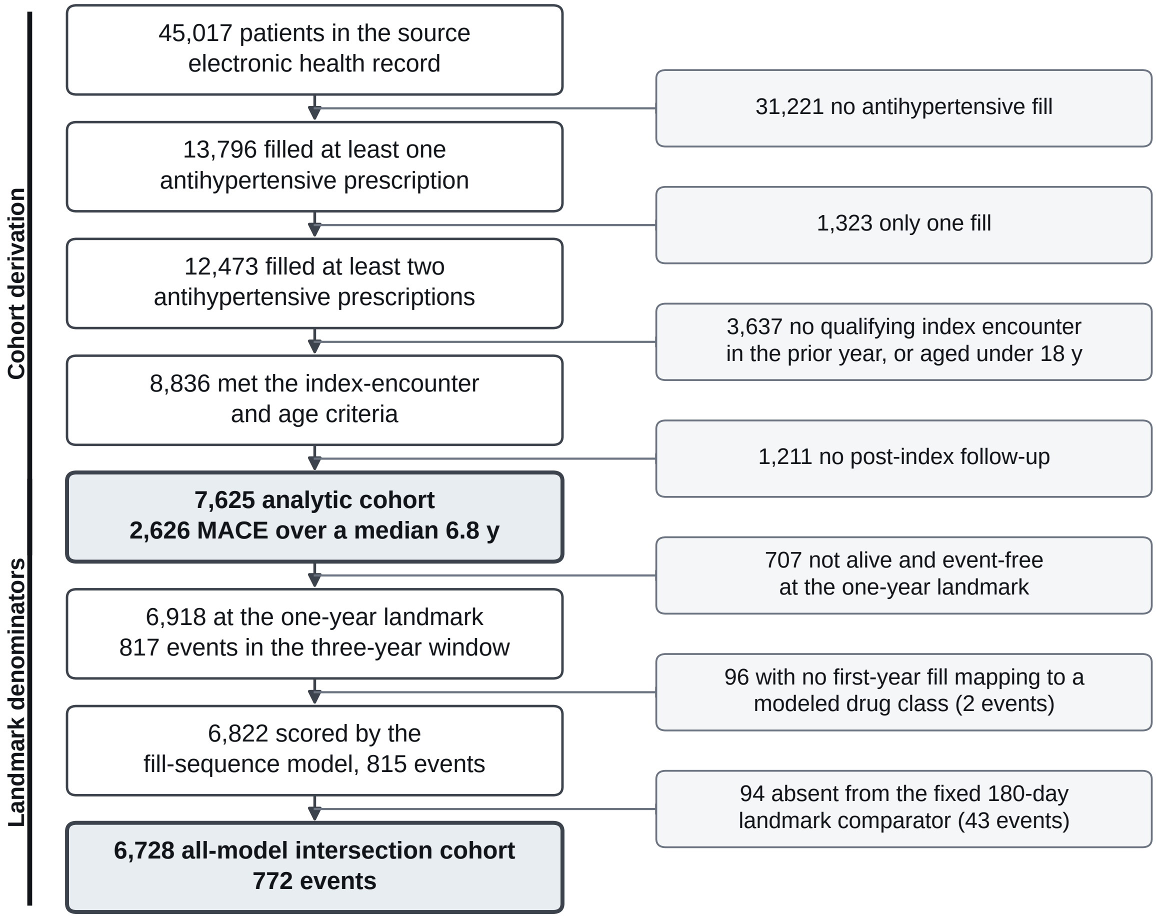

*Notes.* Derivation of the analytic cohort from the source population, and the three strictly nested landmark-stage denominators used in this article. Of 45,017 source patients, 13,796 filled at least one antihypertensive prescription and 12,473 filled at least two; 8,836 of those met the index-encounter and age criteria, and 7,625 met all eligibility criteria and form the analytic cohort. Of the 6,918 patients alive and event-free at the day-365 landmark, 96 cannot be scored by the fill-sequence model, which requires at least one fill in the first year mapping to one of the seven modeled drug classes, leaving 6,822. A further 94 of those are absent from the fixed 180-day landmark comparator, which is defined on a differently constructed eligible population, leaving the 6,728-patient intersection required whenever every model in a comparison must contribute a prediction. Ninety-five patients in total are absent from that comparator, one of whom is also among the 96 above, which is why the two reductions total 190 patients and 45 of the 817 landmark events rather than 191 and 46.

#### Supplementary References

1. Jain S, Wallace BC. Attention is not explanation. In: *Proceedings of the 2019 Conference of the North American Chapter of the Association for Computational Linguistics*. 2019:3543-3556.

2. Vickers AJ, Elkin EB. Decision curve analysis: a novel method for evaluating prediction models. *Med Decis Making.* 2006;26(6):565-574.

3. Pencina MJ, D'Agostino RB Sr, Steyerberg EW. Extensions of net reclassification improvement calculations to measure usefulness of new biomarkers. *Stat Med.* 2011;30(1):11-21.

4. Khan SS, Matsushita K, Sang Y, et al. Development and validation of the American Heart Association's Predicting Risk of Cardiovascular Disease EVENTs (PREVENT) equations. *Circulation.* 2024;149(6):430-449.

5. Efron B. The efficiency of Cox's likelihood function for censored data. *J Am Stat Assoc.* 1977;72(359):557-565.

6. Inker LA, Eneanya ND, Coresh J, et al. New creatinine- and cystatin C-based equations to estimate GFR without race. *N Engl J Med.* 2021;385(19):1737-1749.

7. Markus AF, Verhamme KMC, Kors JA, Rijnbeek PR. TreatmentPatterns: an R package to facilitate the standardized development and analysis of treatment patterns across disease domains. *Comput Methods Programs Biomed.* 2022;225:107081.

8. Antolini L, Boracchi P, Biganzoli E. A time-dependent discrimination index for survival data. *Stat Med.* 2005;24(24):3927-3944.

9. Collins GS, Moons KGM, Dhiman P, et al. TRIPOD+AI statement: updated guidance for reporting clinical prediction models that use regression or machine learning methods. *BMJ.* 2024;385:e078378.

10. Therneau TM, Grambsch PM. *Modeling Survival Data: Extending the Cox Model.* New York, NY: Springer;

2000.

11. Riley RD, Snell KIE, Ensor J, et al. Minimum sample size for developing a multivariable prediction model: PART II - binary and time-to-event outcomes. *Stat Med.* 2019;38(7):1276-1296.

12. Peduzzi P, Concato J, Feinstein AR, Holford TR. Importance of events per independent variable in proportional hazards regression analysis. II. Accuracy and precision of regression estimates. *J Clin Epidemiol.* 1996;49(12):1373-1379.

#### Data and code availability

The analysis code is available from the corresponding author on reasonable request and will be released in a public repository with a citable digital object identifier on acceptance. The patient-level electronic health record and pharmacy dispensing data cannot be shared, owing to privacy and institutional data-use restrictions. All values reported in these materials are aggregate summaries and contain no protected health information.

#### TRIPOD+AI checklist

Completed checklist for Collins GS, Moons KGM, Dhiman P, et al. TRIPOD+AI statement: updated guidance for reporting clinical prediction models that use regression or machine learning methods. *BMJ* 2024;385:e078378. Locations are given by section and subsection rather than page number. Items that do not apply are marked n/a with the reason; items only partly addressed are marked partial.

| Section | Item | Checklist item | Where reported |
|---|---|---|---|
| **TITLE** | | | |
| Title | 1 | Identify the study as developing or evaluating the performance of a multivariable prediction model, the target population, and the outcome to be predicted | Title. Names the target population (antihypertensive medication users) and the outcome domain (cardiovascular risk). **Partial:** the title does not use the word ”development”; the Abstract and Objectives make the design explicit. |
| **ABSTRACT** | | | |
| Abstract | 2 | See TRIPOD+AI for Abstracts checklist | Abstract |
| **INTRODUCTION** | | | |
| Background | 3a | Explain the healthcare context (including whether diagnostic or prognostic) and rationale for developing or evaluating the prediction model, including references to existing models | Background and Significance, paragraphs 1 and 2. Existing models named: Pooled Cohort Equations, PREVENT, and the PDC measure used in Medicare Star Ratings. |
| Background | 3b | Describe the target population and the intended purpose of the prediction model in the context of the care pathway, including its intended users | Background and Significance; Methods, Study population. Intended purpose is stated in the Objective and the Conclusion (complementing existing cardiovascular risk models and population-health workflows). The Discussion states that the sequence model is an existence proof rather than a proposed clinical instrument. |

*continued on next page*

| Section | Item | Checklist item | Where reported |
|---|---|---|---|
| Background | 3c | Describe any known health inequalities between sociodemographic groups | Background and Significance, paragraph 2 (PDC-measured antihypertensive nonadherence is more common among Black and Hispanic patients and in lower-income groups, with reference). The construct is handled in Methods, Study population (race as a sociopolitical category, retained so its contribution can be measured), Methods, Secondary and sensitivity analyses, Results, Temporal and subgroup robustness, and SI Table S9. |
| Objectives | 4 | Specify the study objectives, including whether the study describes the development or validation of a prediction model (or both) | Objective. Development with internal validation; no external validation is claimed. |
| **METHODS** | | | |
| Data | 5a | Describe the sources of data separately for the development and evaluation datasets, the rationale for using these data, and representativeness of the data | Methods, Study design and setting. Michigan Medicine electronic health records linked to outpatient dispensing records from Surescripts. Representativeness is addressed in the Discussion, limitations paragraph. |
| Data | 5b | Specify the dates of the collected participant data, including start and end of participant accrual; and, if applicable, end of follow-up | Methods, Study population. Accrual June 2010 to August 2019; health-system follow-up to May 2025; deaths ascertained through November 2025. |
| Participants | 6a | Specify key elements of the study setting including the number and location of centres | Methods, Study design and setting. One academic health system, Michigan Medicine, Ann Arbor, MI, USA. |
| Participants | 6b | Describe the eligibility criteria for study participants | Methods, Study population; Results, Study cohort; SI Figure S4 (flow diagram). |
| Participants | 6c | Give details of any treatments received, and how they were handled during model development or evaluation, if relevant | Methods, Study population (seven antihypertensive classes) and Representations of the medication history; SI, S-Methods 1 (combination products decomposed to component classes). Treatment history is the exposure of interest, not a nuisance variable. |

*continued on next page*

| **Section** | **Item** | **Checklist item** | **Where reported** |
|---|---|---|---|
| Data preparation | 7 | Describe any data pre-processing and quality checking, including whether this was similar across relevant sociodemographic groups | Methods, Representations of the medication history (30-day interval scaffold, union-exposure PDC, token construction) and Missing data; SI, S-Methods 1 and 2. **Partial:** pre-processing is identical for all patients by construction, but the manuscript does not report a separate check of pre-processing quality by sociodemographic group. |
| Outcome | 8a | Clearly define the outcome that is being predicted and the time horizon, including how and when assessed, the rationale for choosing this outcome, and whether the method of outcome assessment is consistent across sociodemographic groups | Methods, Outcome. Three-point MACE (first myocardial infarction, stroke, or death from any cause), three-year horizon from a one-year landmark. Rationale for all-cause rather than cardiovascular death is given, together with its cost. Ascertainment is administrative and identical across groups. |
| Outcome | 8b | If outcome assessment requires subjective interpretation, describe the qualifications and demographic characteristics of the outcome assessors | **n/a.** Outcomes are ascertained from coded electronic health record data and the state death index. No human adjudication. |
| Outcome | 8c | Report any actions to blind assessment of the outcome to be predicted | **n/a.** Same reason as 8b; blinding does not apply to automated ascertainment from historical records. |
| Predictors | 9a | Describe the choice of initial predictors and any pre-selection of predictors before model building | Methods, Study population (the 22-covariate specification) and Representations of the medication history; SI, S-Methods 1. Covariates were prespecified from the constituents of published risk calculators; no data-driven pre-selection was performed. |
| Predictors | 9b | Clearly define all predictors, including how and when they were measured | Methods, Study population (most recent value on or before the index date) and Representations of the medication history; SI, S-Methods 1; SI Table S15 (distributions of every engineered and token feature). |
| Predictors | 9c | If predictor measurement requires subjective interpretation, describe the qualifications and demographic characteristics of the predictor assessors | **n/a.** All predictors are recorded values or quantities derived from dispensing records. |

*continued on next page*

| Section | Item | Checklist item | Where reported |
|---|---|---|---|
| Sample size | 10 | Explain how the study size was arrived at, and justify that the study size was sufficient. Include details of any sample size calculation | SI, S-Methods 6. No a priori calculation; all eligible patients were analysed. 817 events at the landmark, approximately 21 per candidate predictor in the largest regression model, against the conventional minimum of 10. A prespecified learning-curve analysis assesses data adequacy for the neural model (SI Figure S3). |
| Missing data | 11 | Describe how missing data were handled. Provide reasons for omitting any data | Methods, Missing data; SI, S-Methods 3. Multiple imputation with 20 datasets (miceforest, LightGBM engine, five iterations, seed 42); smoking and race retained as explicit categories. Complete-case sensitivity analysis in SI Table S20. The limitation of full-cohort imputation is disclosed. |
| Analytical methods | 12a | Describe how the data were used in the analysis, including whether the data were partitioned | Methods, Statistical analysis. Five-fold cross-validation repeated across 20 imputed datasets; all reported metrics derive from held-out predictions. |
| Analytical methods | 12b | Depending on the type of model, describe how predictors were handled in the analyses | Methods, Representations of the medication history; SI, S-Methods 2 (log transforms of time, gap and days supplied within the token; dose intensity normalised to the maximum beneficial dose). Collinearity among engineered features is reported in SI Table S13. |
| Analytical methods | 12c | Specify the type of model, rationale, all model building steps, including any hyperparameter tuning, and method for internal validation | Methods, Representations of the medication history; SI, S-Methods 2 (bidirectional LSTM, hidden size 96, attention pooling, two-layer perceptron, Cox partial likelihood with Efron's approximation, AdamW, cosine schedule, early stopping, 20 × 5 × 3 = 300 fits) and Statistical analysis (Cox models, five-fold cross-validation). Rationale for a recurrent rather than transformer architecture is given in SI, S-Methods 2. Software: Statistical analysis names Python with lifelines, XGBoost, PyTorch, scikit-learn and miceforest; package versions are recorded in the environment specification included with the analysis code. |

*continued on next page*

| Section | Item | Checklist item | Where reported |
|---|---|---|---|
| Analytical methods | 12d | Describe if and how any heterogeneity in estimates of model parameter values and model performance was handled and quantified across clusters | **n/a.** Single centre, so no clustering by site. Heterogeneity over calendar time is examined instead (Methods, Secondary and sensitivity analyses; SI Tables S7 and S21). |
| Analytical methods | 12e | Specify all measures and plots used (and their rationale) to evaluate model performance and, if relevant, to compare multiple models | Methods, Statistical analysis. Primary: landmark time-dependent concordance. Prespecified secondary: Antolini's time-dependent concordance. Also calibration, decision-curve net benefit at 7.5%, 15% and 25%, and, in SI, S-Methods 7, integrated discrimination improvement and net reclassification improvement. Paired bootstrap on a common denominator for all model comparisons. |
| Analytical methods | 12f | Describe any model updating arising from the model evaluation, either overall or for particular sociodemographic groups or settings | Methods, Statistical analysis; SI, S-Methods 5. Prespecified recalibration of the neural model by leave-one-fold-out fitting; an overall scaling correction and a flexible monotonic correction were compared. SI Table S6. |
| Analytical methods | 12g | For model evaluation, describe how the model predictions were calculated | Methods, Statistical analysis; SI, S-Methods 4 (held-out linear predictors, Kaplan-Meier calibration of the centred linear predictor to recover absolute risk) and Code availability. |
| Class imbalance | 13 | If class imbalance methods were used, state why and how this was done, and any subsequent methods to recalibrate | **n/a.** No class-imbalance method was used. The models are survival models fitted by partial likelihood, and no resampling, reweighting or threshold shifting was applied. |
| Fairness | 14 | Describe any approaches that were used to address model fairness and their rationale | Methods, Study population (rationale for retaining race as a covariate rather than removing it) and Secondary and sensitivity analyses (prespecified fairness audit across race, sex, age and diabetes status). |

*continued on next page*

| Section | Item | Checklist item | Where reported |
|---|---|---|---|
| Model output | 15 | Specify the output of the prediction model. Provide details and rationale for any classification and how the thresholds were identified | Methods, Statistical analysis. Output is a continuous risk score, converted to an absolute three-year probability by calibration to the observed survival function. No classification threshold is used for the primary analysis; the 7.5%, 15% and 25% thresholds in the decision-curve analysis are taken from the ACC/AHA primary-prevention guideline (reference 1 of the main text). |
| Training versus evaluation | 16 | Identify any differences between the development and evaluation data in healthcare setting, eligibility criteria, outcome, and predictors | **n/a for external evaluation** (none performed). Differences between the internally fitted reference and the externally derived PREVENT equation, which are material to how the anchored ladder should be read, are set out in Methods, Anchoring the ladder on the PREVENT equation. |
| Ethical approval | 17 | Name the institutional research board or ethics committee that approved the study and describe the participant informed consent or the ethics committee waiver of informed consent | Methods, Study design and setting. University of Michigan Medical School Institutional Review Board (IRBMED), HUM00244597; requirement for informed consent waived. |
| **OPEN SCIENCE** | | | |
| Funding | 18a | Give the source of funding and the role of the funders for the present study | Funding. Y.Z. was supported by a Rackham Predoctoral Fellowship, University of Michigan; the funder had no role in the design or conduct of the study, the interpretation of the results, or the decision to submit. |
| Conflicts of interest | 18b | Declare any conflicts of interest and financial disclosures for all authors | Competing interests. Declared per author. |
| Protocol | 18c | Indicate where the study protocol can be accessed or state that a protocol was not prepared | Methods, Study design and setting. The analysis plan was registered internally before model fitting; it is not publicly posted. |
| Registration | 18d | Provide registration information for the study, including register name and registration number, or state that the study was not registered | **Not registered.** The study was not registered in a public register. Stated here rather than in the manuscript. |
| Data sharing | 18e | Provide details of the availability of the study data | Data availability. |
| Code sharing | 18f | Provide details of the availability of the analytical code | Code availability. Available to editors and reviewers on request during peer review; public deposit with a citable DOI on acceptance. |

*continued on next page*

| Section | Item | Checklist item | Where reported |
|---|---|---|---|
| **PATIENT AND PUBLIC INVOLVEMENT** | | | |
| Patient and public involvement | 19 | Provide details of any patient and public involvement during the design, conduct, reporting, interpretation, or dissemination of the study or state no involvement | **No involvement.** No patients or members of the public were involved in the design, conduct, reporting, interpretation, or dissemination of the study. Stated here rather than in the manuscript. |
| **RESULTS** | | | |
| Participants | 20a | Describe the flow of participants through the study, including the number of participants with and without the outcome and, if applicable, a summary of the follow-up time. A diagram may be helpful | Results, Study cohort; SI Figure S4 (flow diagram, including the three nested landmark denominators and their event counts). Median follow-up 6.8 years. |
| Participants | 20b | Report the characteristics overall and, where applicable, for each data source or setting, including key dates, key predictors, treatments received, sample size, number of outcome events, follow-up time, and amount of missing data. Report any differences across key demographic groups | Table 1; SI Tables S1 and S15. Missingness reported per variable in Table 1. Differences by MACE status reported as standardized mean differences; differences by race, sex, age and diabetes reported in SI Table S9. |
| Participants | 20c | For model evaluation, show a comparison with the development data of the distribution of important predictors | **n/a.** No separate external evaluation dataset. The temporal-validation split is described in Methods, Secondary and sensitivity analyses, and SI, S-Methods 7, with split balance in SI Table S21 Panel A. |
| Model development | 21 | Specify the number of participants and outcome events in each analysis | Results, throughout; Discussion, limitations paragraph (190 of 6,918 eligible patients and 45 of 817 events excluded from the paired comparisons); SI, S-Methods 8 (the three nested denominators 6,918 / 6,822 / 6,728); SI Figure S4; and the legend of every supplementary table, which states its denominator and event count (817 / 815 / 772 in the three-year window). |

*continued on next page*

| Section | Item | Checklist item | Where reported |
|---|---|---|---|
| Model specification | 22 | Provide details of the full prediction model to allow predictions in new individuals and to enable third party evaluation and implementation, including any restrictions to access or reuse | The eight-variable parsimonious score is fully specified with coefficients in SI Table S10. The architecture, token schema and training procedure of the fill-sequence model are given in Methods, Representations of the medication history, and SI, S-Methods 2; Code availability states that the code and the trained weights will be deposited in a public archive with a DOI on acceptance. The paper does not propose the neural model for deployment. |
| Model performance | 23a | Report model performance estimates with confidence intervals, including for any key subgroups. Consider plots to aid presentation | Results, all sections; Figures 2 to 6; SI Tables S2, S3, S5, S8, S9, S18, S19. Subgroup estimates in SI Table S9 are reported without intervals, with the reason stated (event counts as low as 26). |
| Model performance | 23b | If examined, report results of any heterogeneity in model performance across clusters | **n/a.** Single centre. Temporal heterogeneity is reported in Results, Temporal and subgroup robustness, and SI Tables S7 and S21. |
| Model updating | 24 | Report the results from any model updating, including the updated model and subsequent performance | Results, Calibration and clinical utility; Figure 4; SI Table S6 (calibration slope, decile gap, integrated Brier score, net benefit and reclassification before and after each correction). |
| **DISCUSSION** | | | |
| Interpretation | 25 | Give an overall interpretation of the main results, including issues of fairness in the context of the objectives and previous studies | Discussion, paragraphs 1 to 7. Fairness is interpreted in paragraph 7, on cross-group discrimination and calibration. |
| Limitations | 26 | Discuss any limitations of the study and their effects on any biases, statistical uncertainty, and generalisability | Discussion, limitations paragraph. Covers single-centre design, model-class confounding of the sequence-versus-summary comparison, the intersection-cohort restriction, and the imputation model. The all-cause death component of the outcome is addressed in Methods, Outcome, and Discussion, paragraph 3 (cause-specific analysis). |

*continued on next page*

| Section | Item | Checklist item | Where reported |
|---|---|---|---|
| Usability | 27a | Describe how poor quality or unavailable input data should be assessed and handled when implementing the prediction model | **Partial.** Results, Risk stratification when laboratory data are unavailable, reports performance under restricted input sets, including a scenario with no laboratory values. Handling of individually missing or poor-quality inputs at the point of use is not specified, because no model is proposed for deployment. |
| Usability | 27b | Specify whether users will be required to interact in the handling of the input data or use of the model, and what level of expertise is required of users | **n/a with reason.** No model is proposed for clinical deployment; the Discussion states that the sequence model is used as an existence proof rather than as a proposed clinical instrument. |
| Usability | 27c | Discuss any next steps for future research, with a specific view to applicability and generalisability of the model | Conclusion. Independent multi-site validation, prospective evaluation of clinical and population-health impact, and assessment of how fill-history representations can complement existing cardiovascular risk models and population-health workflows. |

#### Summary of items not fully met

The items below are collected in one place so that the gaps are visible together rather than distributed through the table above. Each is stated with its reason.

| Item | Status | Reason |
|---|---|---|
| 1 | Partial | The title does not state that a model was developed. |
| 7 | Partial | Pre-processing is uniform by construction but no group-specific quality check is reported. |
| 8b, 8c, 9c | n/a | Automated ascertainment from coded records; no human assessors to characterise or blind. |
| 12d, 20c, 23b | n/a | Single centre and no external evaluation dataset. |
| 13 | n/a | No class-imbalance method used. |
| 16 | n/a | No external evaluation; the PREVENT comparison is described instead. |
| 18d | Not registered | The study was not entered in a public register. |
| 27a | Partial | Restricted-input performance is reported; point-of-use handling is not, because no model is proposed for deployment. |
| 27b | n/a | No model proposed for deployment. |
